# Immunologic and epidemiologic drivers of norovirus transmission in daycare and school outbreaks

**DOI:** 10.1101/2019.12.19.19015396

**Authors:** Joshua Havumaki, Joseph NS Eisenberg, Claire P Mattison, Benjamin A Lopman, Ismael R Ortega-Sanchez, Aron J Hall, David W Hutton, Marisa C Eisenberg

**Affiliations:** Department of Epidemiology; University of Michigan; National Center for Immunization and Respiratory Diseases, Division of Viral Diseases; Centers for Disease Control and Prevention; Oak Ridge Institute for Science and Education; Department of Epidemiology, Emory University; Departments of Health Management and Policy, Industrial and Operations Engineering; University of Michigan; Departments of Epidemiology, Mathematics, Complex Systems; University of Michigan

**Keywords:** norovirus, transmission model, children, surveillance data

## Abstract

1

**Background:** Norovirus outbreaks are notoriously explosive, with dramatic symptomology and rapid disease spread. Children are particularly vulnerable to infection and drive norovirus transmission due to their high contact rates with each other and the environment. Despite the explosive nature of norovirus outbreaks, attack rates in schools and daycares remain low with the majority of students not reporting symptoms.

**Methods:** We explore immunologic and epidemiologic mechanisms that may underlie epidemic norovirus transmission dynamics using a disease transmission model. Towards this end, we compared different model scenarios, including innate resistance and acquired immunity (collectively denoted ‘immunity’), stochastic extinction, and an individual exclusion intervention. We calibrated our model to daycare and school outbreaks from national surveillance data.

**Results:** Recreating the low attack rates observed in daycare and school outbreaks required a model with immunity. However, immunity alone resulted in shorter duration outbreaks than what was observed. The addition of individual exclusion (to the immunity model) extended outbreak durations by reducing the amount of time that symptomatic people contribute to transmission. Including both immunity and individual exclusion mechanisms resulted in simulations where both attack rates and outbreak durations were consistent with surveillance data.

**Conclusions:** The epidemiology of norovirus outbreaks in daycare and school settings cannot be well described by a simple transmission model in which all individuals start as fully susceptible. Interventions should leverage population immunity and encourage more rigorous individual exclusion to improve venue-level control measures.

## 3 Background

Norovirus is the leading cause of acute gastroenteritis across all ages in the United States (US), with 19 to 21 million cases occurring per year [1]. The role of children in its transmission has recently been highlighted in a mathematical modeling analysis [2], which found that pediatric vaccination would result in substantially higher protective population-level effects when compared with vaccination of the elderly. This finding highlights the propensity of children in daycares and schools to cause explosive outbreaks and subsequently to propagate disease into the community through person-to-person transmission [3–10]. Here we examine how norovirus spreads within schools and daycare centers to inform the potential design of effective venue-specific interventions to ultimately reduce population-level risk.

Norovirus transmission can occur directly through person-to-person contact [11] and indirectly through water [12], food [13], or fomite-mediated pathways [14–17]. Symptomatic individuals efficiently spread virus through vomiting and defecation [17]. After symptoms resolve, individuals continue to shed for an average of ∼2 weeks [18]. Norovirus transmission is sustained through the combination of efficient and prolonged human shedding [17], and extended environmental persistence [19–22]. Additionally, norovirus is highly infectious, with an infectious dose of 18-2800 virions being sufficient to cause infection, while peak viral concentration per gram of stool reach levels of 10^9^ [23, 24]. These features of transmission as well as a lack of long-lasting immunity in human hosts [25, 26], contribute to venue-level norovirus outbreaks potentially exhibiting rapid, explosive growth rates [27, 28]. These explosive epidemic growth rates would be expected to correspond to high attack rates with exhaustion of susceptibles, similar to other highly transmissible diseases like measles [29, 30]. However, despite this explosive tendency and the important role that children play in transmission, attack rates (ARs) in daycare and school outbreaks are relatively low (∼22% to ∼15% in daycares and schools, respectively, based on data from the National Outbreak Reporting System (NORS) [31]).

While the transmission of other explosive diseases, such as measles, may be often be captured through simple susceptible-infectious-recovered (SIR) models in which all individuals start as fully susceptible, norovirus does not result in exhaustion of susceptibles, suggesting that a more complex transmission process may be occurring. One distinction between norovirus and measles is that norovirus immunity is strain dependant and not permanent, so that norovirus outbreaks often occur in populations with immunity. This raises the question of whether population-level immunity is sufficient to explain the norovirus outbreak patterns that we observe.

There are multiple explanations for the combination of explosive outbreaks and low ARs observed in outbreak data. First, ∼20% of the US population lack a functional FUT2 gene, conferring innate resistance to certain norovirus genotypes [32]. Furthermore, depending on age, up to ∼90% of children *<* 5 years of age have norovirus antibodies titers potentially indicating acquired immunity [33, 34], although the level of protection conferred by these antibodies is not known and the levels of acquired immunity may fluctuate in part due to the genetic drift of norovirus strains [35]. Second, the Centers for Disease Control and Prevention (CDC) recommends various interventions to prevent and control norovirus outbreaks, including isolation of individuals during and 1 to 3 days following the symptomatic period [36] which may also reduce transmission [37]. Finally, stochastic extinction may lead to outbreaks ending without exhaustion of susceptibles, especially for smaller populations [38] e.g. daycares. Any combination of these factors may contribute to low ARs within venues.

In this paper, we employ mathematical models to explore underlying mechanisms leading to disease transmission dynamics that can explain the observed norovirus epidemiology within daycare and school venues. Given the epidemiological features discussed above, explaining norovirus dynamics requires a detailed representation of the mechanisms driving transmission. Here, we examine which mechanisms are sufficient to explain the epidemiological patterns seen in outbreaks using a transmission model calibrated to CDC NORS surveillance data.

## 4 Methods

## 5 NORS Dataset

We calibrated our model to outbreak duration (in days), student AR, and student population size data from NORS, a CDC-managed internet-based surveillance system through which state, territorial, and local health departments within the US can enter outbreak information [31, 39] (See Appendix Table S6 for summary statistics of dataset). Our dataset includes all school and daycare outbreaks in NORS that occurred from 2009–2016 which indicated norovirus as the only suspected or confirmed etiology. We classified a given venue as daycare or school based on self-reported classification by the reporting agency [40]. In total, there were 989 school outbreaks and 329 daycare outbreaks, which comprised 4.6% of all outbreaks reported through NORS during 2009–2016 (i.e., 1,318 of 28,580 total outbreaks across all modes and etiologies). We only included outbreaks with complete data (i.e., not missing for total students exposed, AR, and outbreak start and end dates), and with ARs and durations within the 5^*th*^ and 95^*th*^ percentiles of the dataset to remove outliers and to calibrate our model to data generally representative of common norovirus outbreaks. Our final calibration dataset consisted of 562 total norovirus outbreaks.

### 5.1 Model Structure

We modeled transmission in school and daycare venues and examined how including different mechanisms capture the features of the NORS data. Our stochastic model is an extension of [2]. All analyses were conducted in R version 3.2.4 [41].

#### 5.1.1. Transmission Model for Daycare centers and Schools

Transmission occurs directly through person-to-person contact or indirectly through fomite-mediated pathways i.e., shedding and pickup of virions in the environment (in this analysis, we are not simulating foodborne or waterborne transmission). Individuals start as susceptible *S*, partially immune *P*, or have innate resistance (and start as fully recovered) *R* depending on the initial conditions (Figure 1). Susceptible and partially immune individuals become infected according to the force of infection *λ*(*t*), which is derived from the number of symptomatic individuals (*I*), asymptomatic individuals (*A*_1_, *A*_2_, and *A*_3_), environmental pathogens residing on fomites (*F*_1_ and *F*_2_), and the human-to-human (*β*_*HH*_) and fomite-to-human (*β*_*F H*_) transmission rate parameters. The parameter *β*_*A*_ represents the reduction in efficiency of asymptomatic transmission compared with symptomatic transmission. Excluded individuals do not contribute to the force of infection:

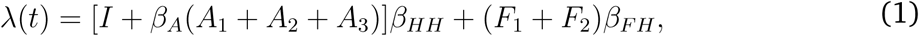

**Figure 1:**
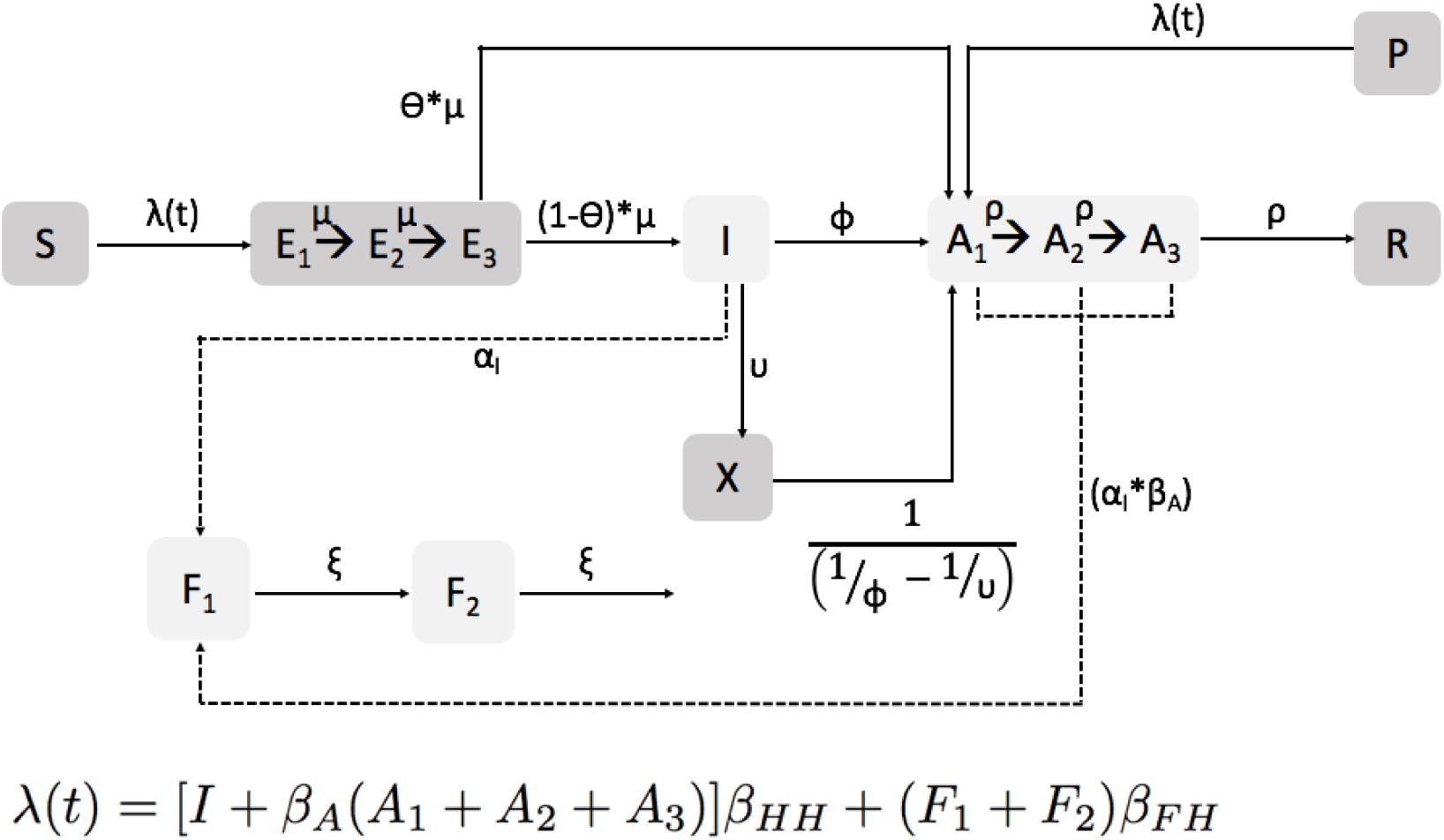
Model schematic: All compartments involved in the force of infection (Equation 1) are in light gray. The full force of infection equation is also shown in the figure above. Susceptible individuals (*S*) may be infected and pass through a latent period (*E*_1_ to *E*_3_) before becoming symptomatically (*I*) or asymptomatically infectious (*A*_1_ to *A*_3_). Once recovered from asymptomatic infection, individuals become fully immune (*R*). Note, there is no waning immunity due to the fact that we are simulating over a short timescale (i.e., 60 days). At the start of the outbreak some individuals may be fully immune (*R*) due to innate resistance or partially immune (*P*) due to acquired immunity. Social distancing or individual exclusion is represented by *X*. During infection, individuals may shed pathogens onto environmental fomites (*F*_1_). As pathogens on the fomites decay, they move to (*F*_2_), together the two fomite compartments represent biphasic decay of norovirus in the environemnt. All parameter values are shown in Table 1.

**Table 1:**
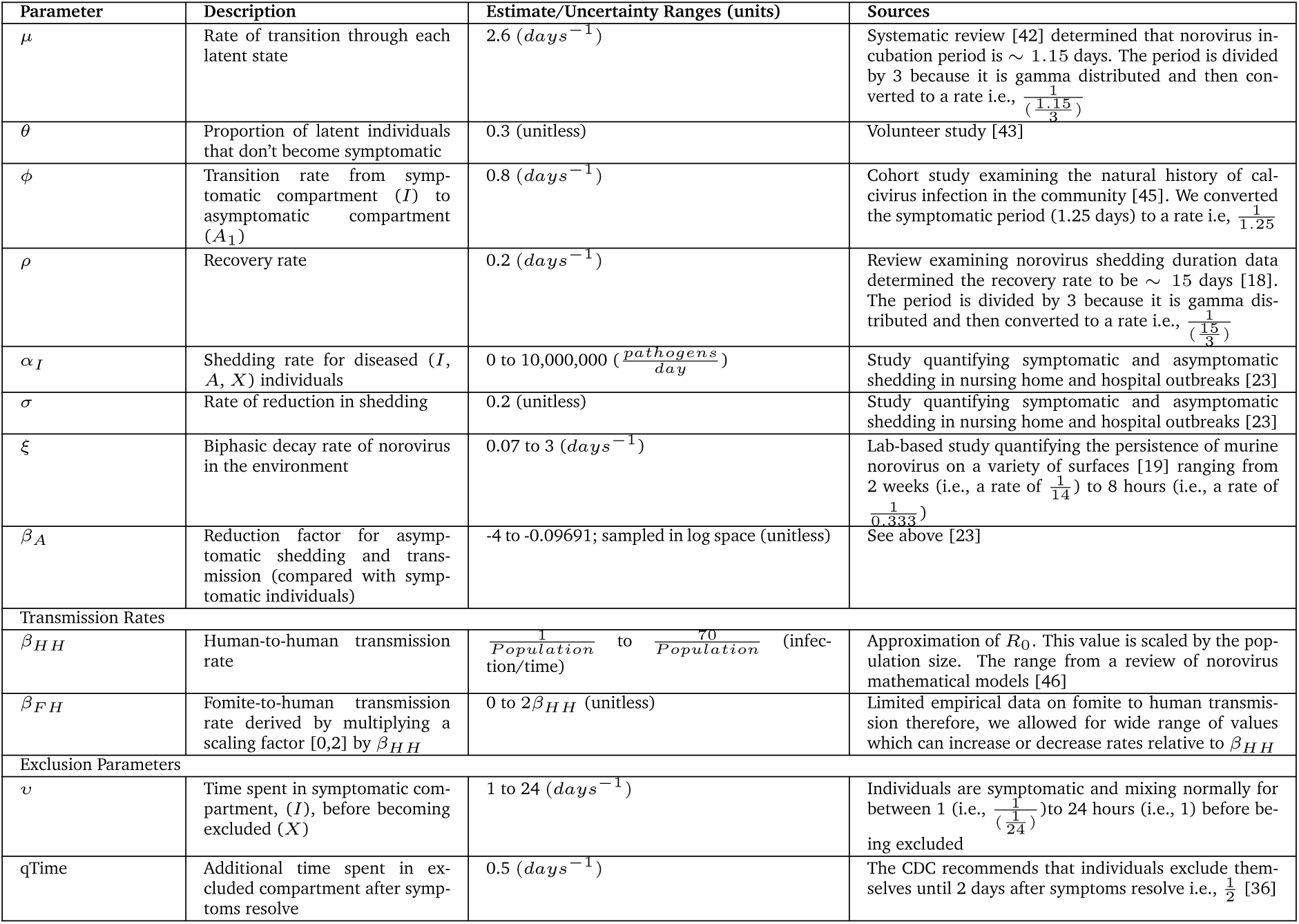
Transmission Model Parameter Values and Uncertainty Ranges

Infected individuals move through a gamma-distributed latent period (*E*_1_, *E*_2_, and *E*_3_) which represents the distribution of the incubation times in the literature [42]. Individuals then become either symptomatic or asymptomatic [43]. The gamma-distributed asymptomatic period represents post-symptomatic shedding and exhibits reduction in shedding by stage (e.g., individuals in *A*_2_ shed less than individuals in *A*_1_). Partially-immune individuals can become infected, but not diseased (symptomatic). Symptomatic individuals may become excluded (*X*) until 2 days after their symptoms resolve and do not contribute to transmission while they are excluded. All non-excluded symptomatic and asymptomatic individuals shed pathogen into the environment (in the venue). Norovirus pathogen decay on fomites occurs in a biphasic pattern with an initial rapid die-off followed by a period of slower decay [19, 44]. Finally, all individuals who become infected eventually progress to fully recovered. See Appendix Table S1 for initial condition ranges, and Section S2 for the model description and equations.

#### 5.1.2. Predefined parameter ranges

To reduce parameter space and the number of simulations required to calibrate the model, we fixed the values of parameters that have been estimated in the literature (i.e., *µ, θ, ϕ, ρ, σ*, and *qTime*). If there was no information about a given parameter (i.e., *α*_*I*_, *ξ, β*_*A*_,*β*_*HH*_, *β*_*F H*_, and *υ*), we sampled its value from predefined ranges. See Table 1 for all parameter values and ranges.

### 5.2 Model Scenarios

We considered the following scenarios to examine mechanisms that can recreate the features of norovirus transmission:

- **Baseline Model**: The outbreak starts in a fully susceptible population.
- **Immunity Model**: The outbreak starts in a population where 20% of individuals have innate resistance [32] and between 0 and 80% of individuals have acquired immunity (depending on the sampled initial condition values, see Section S5). There is no individual exclusion.
- **Individual Exclusion Model**: The outbreak starts in a fully susceptible population. Symptomatic individuals can become excluded (*X*) and these individuals do not contribute to transmission.
- **Combined Model**: The outbreak starts in a population with immunity (i.e., innate resistance and acquired immunity) and contains symptomatic individuals that can become excluded.

See Appendix Section S3 for more details about the model scenarios. All models were simulated in daycares and schools. We randomly sampled starting population sizes from the distribution of exposed student populations in the NORS data (Table S6). To address assumptions about our modeling framework, we conducted sensitivity analyses. First, we looked to see whether specific initial conditions influence our results in our best-calibrated model. We varied whether the outbreak was initiated by an infected individual entering the venue or by differing amounts of contamination in the environmental. Second, we looked to see whether including staff in addition to student transmission influenced our conclusions (see Appendix Sections S11 and S12 for details).

### 5.3 Calibration

We calibrated each venue–specific model separately to its corresponding NORS data using sample-importance-resampling [47]. This approach allowed us to obtain an array of parameter sets that best recreate the NORS data distribution.

Sample-importance-resampling is a three-step procedure. In the ‘sample’ stage we ran the model with 10,000 randomly sampled parameter and initial condition sets (collectively denoted ‘parameter sets’, and listed in Table 1) from pre-defined ranges using Latin Hypercube Sampling [48]. Each parameter set was run in a school setting and separately, in a daycare setting. Next, in the ‘importance’ stage, we calculated a likelihood to determine how well each parameter set corresponds to the NORS data. Our likelihood compared the 3-dimensional probability distribution of the student ARs, student population size, and outbreak durations from the model to NORS data. We also examined the effects of weakening our prior (i.e., our best guess pre-defined parameter ranges) on the results. Finally, in the ‘resampling’ stage we resampled the parameter sets 2,500 times with replacement using the likelihoods as weights to obtain a final array of parameter sets. See Appendix Section S6 for more details on calibration and the effects of weakening our prior.

As described above, we considered two versions of the model and calibration—one model which included only students (using the student AR and population data), and a second sensitivity analysis that included staff members and students separately in the model (see Appendix Section S12 for details). The overall outbreak duration was used for calibration in both model versions. Calibrating the staff and student model was substantially more computationally expensive than the student-only model. Additionally, the full calibration results for the staff and student model were similar to the student-only model (see below and Appendix Section S12), and so for simplicity we present only the student model here and the model with the staff included is presented as a sensitivity analysis in the appendix.

To determine the best-fitting model, we derived a kernal density estimate of the calibrated model results (namely the density of resamples across AR, population size, and outbreak durations), and calculated the Kullback-Leibler divergence to measure the difference between the each calibrated model and NORS kernal density estimate [49]. We also examined pairwise scatter plots of ARs, outbreak durations, and population sizes for resampled model runs and compared them to the NORS data to assess the calibration graphically.

## 6 Results

### 6.1 NORS Data

Our final dataset (after removing incomplete data and outliers) consisted of 165 daycare outbreaks and 397 school outbreaks. The median student population, AR, and outbreak duration for daycare outbreaks were 80 people (range: 7, 410), 21.5% (range: 4.6%, 69.2%), and 13 days (range: 2, 40 days), respectively. The median population, AR, and duration for school outbreaks were 420 people (range: 6, 6486), 15.3% (range: 4.6%, 68.4%) and 8 days (range: 1, 32 days), respectively. See Appendix Table S6 for details.

### 6.2 Calibration

We found that the fully uninformed prior resulted in the best graphical fit and lowest Kullback-Leibler divergence (discussed in Appendix Section S7). The graphical results of the calibration are shown in the Appendix Section S10 and the Kullback-Leibler divergence for each model is given in Table 2.

**Table 2:** Kullback Leibler divergence for each model compared with the NORS data kernel density estimated distribution. Smaller Kullback-Leibler divergence indicates a more similar distribution to NORS (i.e. less information difference between the NORS and the model distribution). The combined model is shown in bold. The sensitivity analyses include varying the environmental contamination at the start of the outbreak (‘Seeding: Varying Pathogens in Environment’), and seeding with a single diseased individual (‘Seeding: Diseased Individual’)

| Model | Daycare | School |
| --- | --- | --- |
| Stochastic Simulations |  |  |
| Baseline | 5.05 | 7.41 |
| Immunity | 1.05 | 0.53 |
| Individual Exclusion | 1.39 | 3.59 |
| **Combined** | **0.3** | **0.39** |
| Sensitivity Analyses on Combined Model |  |  |
| Seeding: Varying Pathogens in Environment | 0.56 | 0.35 |
| Seeding: Diseased Individual | 0.34 | 0.46 |

### 6.3 Model Comparisons

Figure 2 shows the median attack rates and durations for the NORS data and each of the models. Overall, the combined model matched the NORS data best both graphically and according to Kullback-Leibler divergence (see Figure 3 and Appendix Figures S10 for scatter plots showing model results compared with NORS data and Table 2 for all KullbackLeibler divergences). In the combined model, the daycare and school-aged ARs had medians of 20% (CI: 2.6% to 57.1%) and 14.3% (95% CI: 2.1% to 52.7%), respectively. These are slightly lower, but relatively consistent with the NORS ARs which were 21.5% (range: 4.6% to 69.2%) and 15.3% (range: 4.6% to 68.4%) for daycares and schools, respectively. The daycare and school outbreak durations were 12 days (95% CI: 2 to 36) and 10 days (95% CI: 3 to 31), respectively. These ranges are somewhat smaller, but generally consistent with the NORS outbreak durations which were 13 days (range: 2 to 40) and 8 days (range: 1 to 32) for daycares and schools, respectively. Results for the other models that did not calibrate as well are shown in Tables 3 and 4.

**Table 3:** Venue-specific Attack Rates for All Models: Median (95% CI) [Mean]

| Model | Daycare | School |
| --- | --- | --- |
| Baseline | 57.1% (2.8%, 73.3%) [47%] | 56.5% (0.6%, 70.8%) [48.1%] |
| Immunity | 19.5% (3.1%, 55.1%) [22.2%] | 13.7% (2.6%, 54.2%) [18.3%] |
| Individual Exclusion | 27.3% (2.8%, 70%) [31.1%] | 12.5% (0.7%, 67.8%) [22.8%] |
| Combined | 20% (2.6%, 57.1%) [22.8%] | 14.3% (2.1%, 52.7%) [18.6%] |
| **NORS** | 21.5% (4.6%, 69.2%) [25.3%] | 15.3% (4.6%, 68.4%) [20.4%] |

**Table 4:** Venue-specific Outbreak Durations for All Models: Median (95% CI) [Mean]

| Model | Daycare | School |
| --- | --- | --- |
| Baseline | 8 days (2, 22) [8.8] | 5 days (2, 17) [6.2] |
| Immunity | 8 days (2, 26) [9.7] | 8 days (2, 27) [9.8] |
| Individual Exclusion | 10 days (3, 31) [11.8] | 9 days (2, 30) [11.2] |
| Combined | 12 days (2, 36) [13.5] | 10 days (3, 31) [12] |
| **NORS** | 13 days (2, 40) [14.6] | 8 days (1, 32) [10.8] |

**Figure 2:**
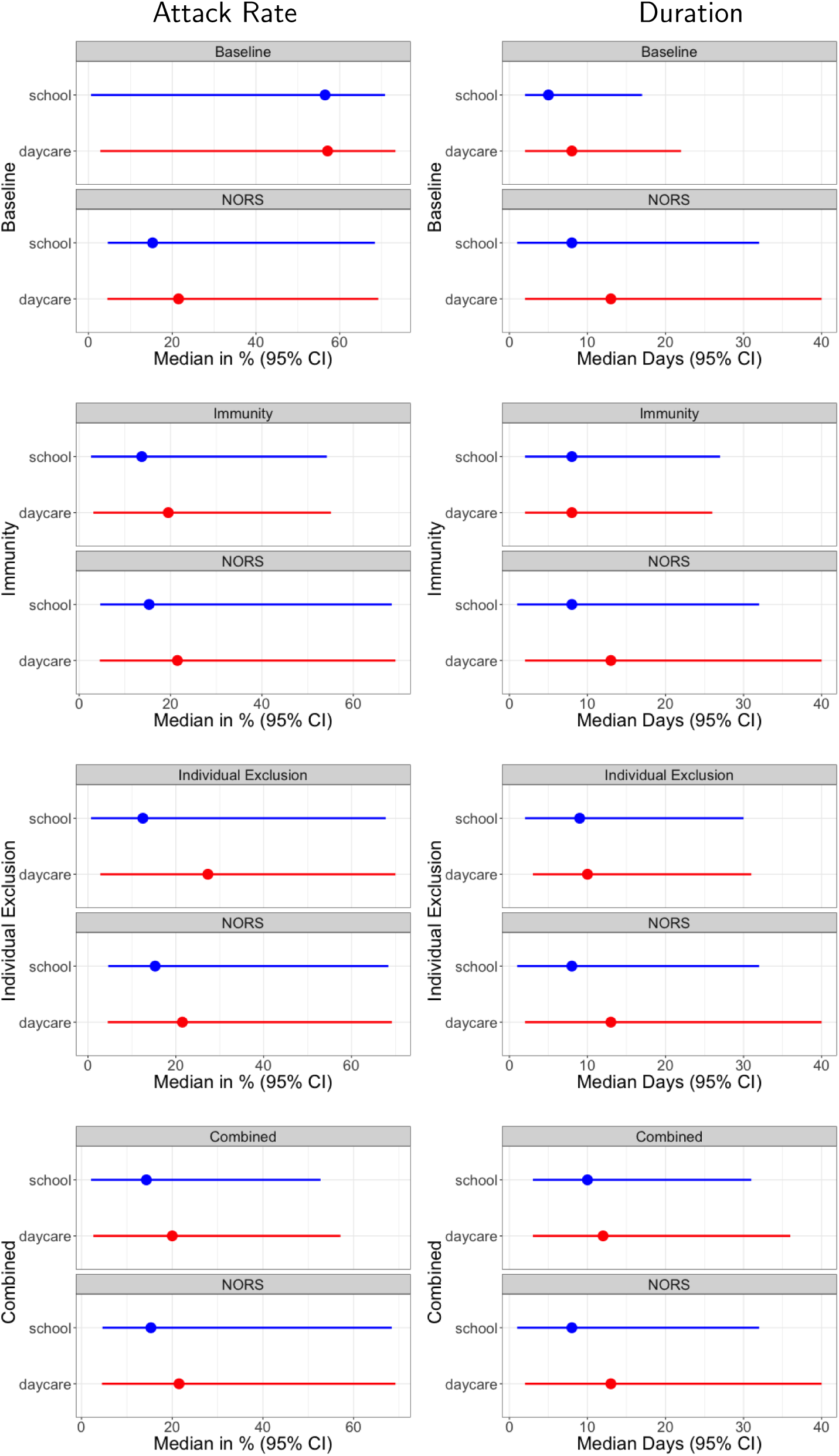
ARs (left column) and durations (right column) for each model compared with NORS data. Each plot corresponds to a different model structure (indicated on the y-axis label).

**Figure 3:**
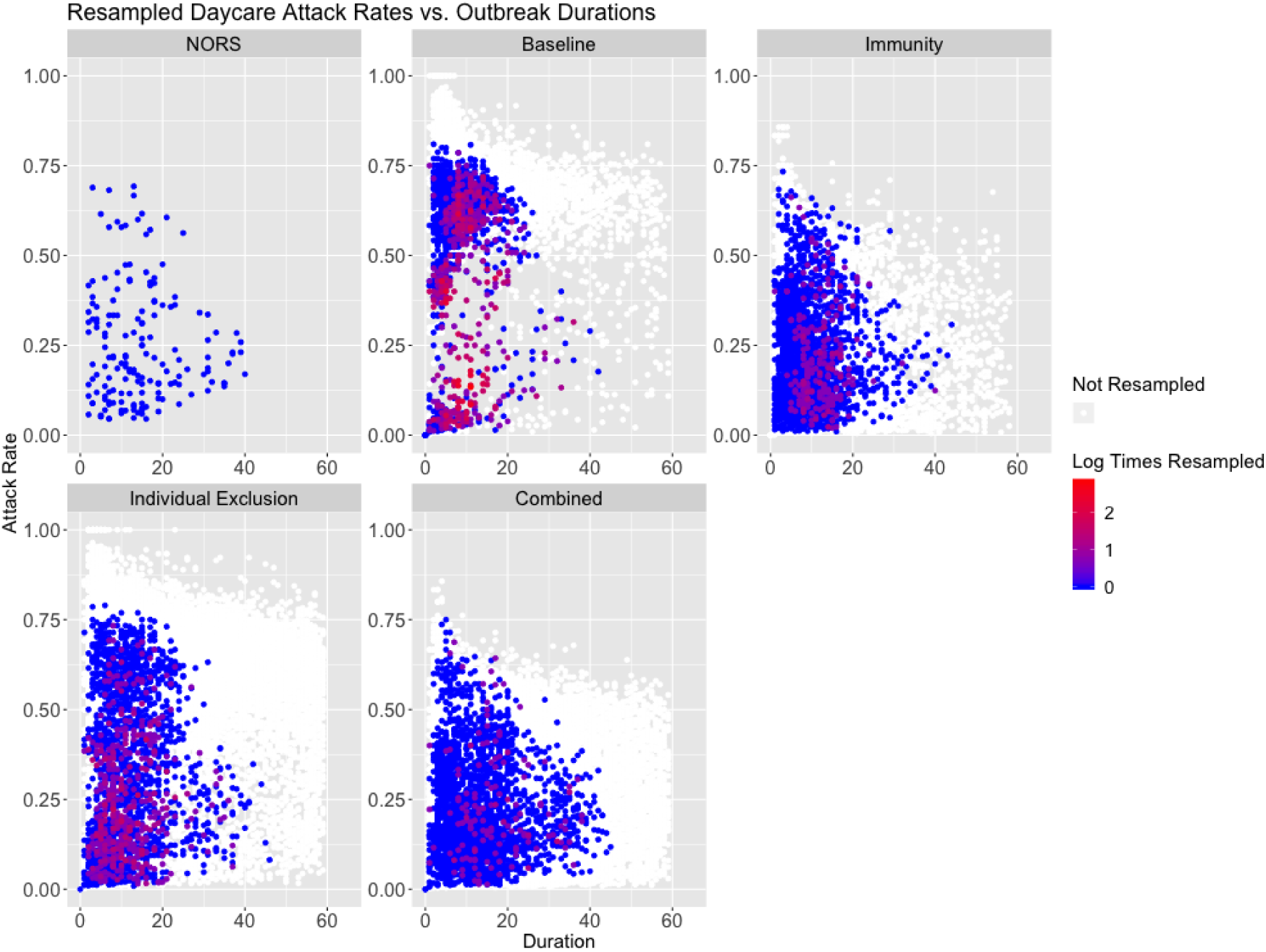
Attack rate vs. outbreak duration in daycares for the NORS data (top left), with all remaining panels showing results from resampled parameter and initial conditions by model scenario. Points correspond to individual parameter sets and are colored by the amount of times they were resampled.

Overall, the baseline model which examined the effects of stochasticity alone (i.e., in the absence of immunity and individual exclusion) resulted in ARs being too high and outbreak durations being too short compared with the NORS data. Among the mechanisms we examined, population immunity (included in the immunity and combined models) was best able to recreate relatively low ARs observed in the NORS data (Table 3). This is because individuals who are partially immune may become infected, but not symptomatic. Even though these individuals contribute to transmission, they are not detected as diseased and do not count in the overall AR. On the other hand, the individual exclusion mechanism (included in the individual exclusion and combined models) resulted in durations that were more consistent with NORS data. Specifically, individual exclusion led to longer outbreak durations (see Appendix Table 4) indicating that it may slow the spread of norovirus. Excluded individuals transmit for a shorter time while they are symptomatic (i.e., between 1 hour and 1 day before being excluded) compared with non-excluded individuals (i.e., 1.25 days). This reduction in transmission time likely prevents the outbreak from spreading as fast as the baseline or immunity models. Individual exclusion also provides population-level protection from becoming symptomatic (as indicated by the reduction in individual exclusion ARs compared with the baseline model).

Each single mechanism model can account for single aspect of the NORS data. Specifically, immunity can recreate the NORS attack rates and individual exclusion can recreate the NORS outbreak durations. Furthermore, examining the pairwise comparison between attack rates and populations and durations and populations, we can see that the immunity and combined models best match the NORS data. See Appendix Figures S2, S3, S5, and S6 for populations plotted against attack rates and outbreak durations. Because of the balance in fit of these different features, the combined model performed best according to the Kullback-Leibler divergence (see Table 2).

For all models, the median outbreak durations were less than the average time it took the first incident cases to fully recover (i.e., less than the ∼ 16-day infectious period), which meant that outbreaks tended to end within a single infectious period indicating a rapid (explosive) spread of disease within venues. Therefore, both the combined and immunity models provided a mechanism for explosive outbreaks that did not result in the entire population becoming diseased.

### 6.4 Sensitivity Analyses

We examined how results changed when simulating different initial conditions and also, including staff transmission. First, starting with different initial conditions using the combined model (seeding with an infected individual or varying the number of pathogens starting in the environment) led to similar model results compared with the original model. See Appendix Sections S11, and S12 for more details, and Tables S7 and S8 for ARs and durations. Second, the student and staff model had qualitative results consistent with the main analysis. Specifically, among calibrated parameter sets, although median results were closer than the student only model to the NORS data for certain model scenarios (e.g. the attack rates of the baseline model), the overall behavior of each scenario was consistent. This is evidenced by the attack rate ranges being wider for the immunity model, and the outbreak duration ranges being wider for the individual exclusion model. Overall, the student and staff model was consistent with our overall results in that the combined model calibrated best to the NORS data (see Appendix section S12 for details).

## 7 Discussion

Although norovirus is classically characterized by explosive outbreaks, its epidemiology exhibits interesting properties that are not captured by simple SIR models starting with a fully susceptible population. The NORS data has lower attack rates and longer durations than were observed in our baseline model alone. An interesting characteristic of norovirus is that a portion of the population exhibits immunity that provides some degree protection. Immunity to norovirus (either innate or post-exposure) is well known, our analysis suggests that this process may be sufficient to explain the observed general pattern of explosive norovirus outbreaks but relatively low ARs within venues. However, this mechanism alone resulted in outbreak durations that were substantially faster than what we observed in the data. Another interesting feature of norovirus outbreaks, due to the short but intense symptomatic phase, is that individual’s choose to and/or are asked to exclude themselves from the general population. This exclusion appears to slow transmission and result in lower attack rates, but does not produce a wide distribution of attack rates in our model. For this reason, the combined model (with both immunity and individual exclusion) was able to generate a wider distribution of outbreaks with both low attack rates and longer outbreak durations and therefore matched the NORS data most closely. These findings suggest that partial population immunity and exclusion of ill individuals are important mechanisms that contribute to the observed characteristics of reported norovirus outbreaks.

These results are based on an analysis of the NORS dataset in conjunction with a transmission model. Both the data and the model come with assumptions and limitations. With respect to the data, NORS relies on passive surveillance, which means there is under-reporting of outbreaks that likely results in the distribution of ARs and durations not being representative of the general trends in the population [11]. Second, the amount of missing data (only 562 of the initial 1,318 outbreak were complete and therefore included) could result in bias. Third, interventions (e.g. decontamination) were likely implemented in many of the reported outbreaks, but the majority (¿ 90%) did not report on this. Fourth, the exposed population reported (i.e., the denominator of the ARs) may correspond to a single classroom, grade-level or entire school, and is not consistent across outbreaks (and the method of determining the exposed population is not specified). This may result in lower ARs (if the exposed population is overestimated) and could have substantial effects on how well our models calibrate. To address this, in our initial exploration of the NORS data, we plotted ARs vs. duration while stratifying on exposed population size (for populations *<* 200) and found that the overall distribution appeared similar across strata (see Appendix Figure S1). Lastly, classifications of school or daycare venues relied on self-reporting. Many venues have a mixture of different age groups and therefore, classifications were likely not consistent.

Our analytical methods include a wide range of assumptions required to generate a parsimonious model. First, we did not explicitly include the processes of waning immunity and the existence of different norovirus strains because we are simulating a single outbreak. These processes are represented in our model by varying the distribution of individuals who start as immune compared with fully susceptible. Second, we assume that partially immune individuals may become infected, but not diseased. In reality, individuals with acquired immunity may become symptomatic or asymptomatic. However, altering the relative proportion of asymptomatic vs. symptomatic individuals i.e., by varying the initial conditions of those who start as partially immune should account for these effects. Third, we used a compartmental rather than individual-based structure, which does not include explicit contact network effects such as clustering and degree heterogeneity. One form of contact heterogeneity is the different transmission rates for staff and students—while our main model did not include staff transmission, the staff and student sensitivity analysis included these features and had results consistent with the main analysis i.e., immunity allows for a wider range of attack rates and individual exclusion allows for a wider range of outbreak durations. For more details see Appendix Section S12. Finally, we assume that each outbreak is seeded in the environment with a set number of pathogens. We explored variations to this in sensitivity analyses altering initial seeding (1) with a single infected individual and (2) varying the number of pathogens starting in the environment. These additional analyses did not substantially change the results (see Appendix Section S11).

Future analyses should consider how interventions can leverage the fact that innate and partial immunity, as well as individual exclusion may shape outbreak patterns. The importance of innate and acquired immunity in reducing attack rates supports the current approach to implement a vaccine and also may be factored into the design of vaccination programs (because ∼ 20% of the population already has innate immunity). The observation that individual exclusion leads to slower transmission, suggests that other interventions like decontamination can work in conjunction with individual exclusion to stop transmission altogether.

## Data Availability

The surveillance datasets are available from the Centers for Disease Control and Prevention upon request and application. Computing code available from the corresponding author upon reasonable request.

## 8 Acknowledgements

We would like to thank Molly Steele Environmental Health Emory University, Zachary Marsh Centers for Disease Control and Prevention, and Dr. Lisa Prosser Department of Pediatrics and Communicable Diseases at the University of Michigan Medical School for their helpful comments, advice, and support. The findings and conclusions in this report are those of the authors and do not necessarily represent the official position of the Centers for Disease Control and Prevention.

## S1 Supplementary Information

## S2 Transmission Model for Daycare Centers and Schools

Transmission occurs either directly through person-to-person contact or indirectly through fomite-mediated pathways i.e., shedding and pickup of virus in shared environments. Individuals start as either susceptible *S*, partially immune *P*, or fully recovered *R* depending on acquired immunity and innate resistance status (Figure 1). Susceptible and partially immune individuals become infected according to the force of infection *λ*(*t*), which is based on: (1) the number of symptomatic (*I*), and asymptomatic (*A*_1_, *A*_2_, and *A*_3_) individuals; the number of pathogens on fomites in the environment (*F*_1_ and *F*_2_); (3) the human to human and fomite to human transmission rates (*β*_*HH*_; and *β*_*F H*_); and (4) The asymptomatic transmission reduction factor (*β*_*A*_) which reduces the efficiency of transmission compared with symptomatic individuals. Excluded individuals, *X*, do not contribute to transmission.

### Force of Infection

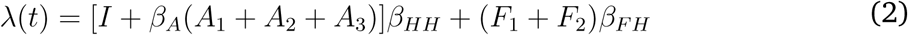

Once infected, individuals pass through a gamma distributed latent period i.e. *E*_1_, *E*_2_, and *E*_3_. It is gamma distributed to represent the empirical distribution of incubation times [42] in the literature. After they pass through the latent period, they become symptomatic. After an individual is symptomatic, they pass through a gamma distributed asymptomatic period i.e., *A*_1_, *A*_2_, and *A*_3_ that represents post-symptomatic shedding and exhibits a reduction in shedding by stage (e.g., individuals in *A*_2_ shed less than individual in *A*_1_, see below for details). A proportion of infected individuals who originate in *S* do not become symptomatic and pass directly from *E*_3_ to *A*_1_. Individuals who start as partially immune can become infected, but do not become symptomatic and move directly to *A*_1_.

A proportion of symptomatic individuals become excluded and move into the *X* compartment. After their symptoms resolve, they move to *A*_1_ and return to the general population with the normal transmission and shedding rates for the *A*_1_ compartment. Finally, all individuals who become infected eventually progress to the fully recovered state. All symptomatic and asymptomatic individuals (unless excluded) shed pathogen into the environment as follows:

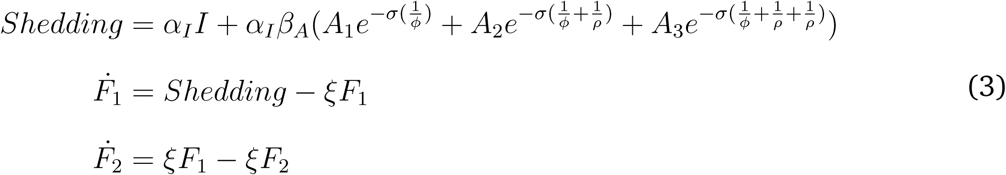

where *α*_*I*_ is the shedding rate for symptomatic individuals, and the reduction factors for shedding among asymptomatic individuals is *β*_*A*_.

The amount of shedding is reduced exponentially as individuals progress across the gamma distributed asymptomatic period by *σ* for each state transition [23]. The symptomatic period, *ϕ*, and the recovery rate, *ρ* (i.e., from *A*_1_ to *A*_2_ etc.), account for the length of time that individuals shed at certain rates.

Viral concentration on fomites is tracked in the venue. Norovirus pathogen decay on fomites occurs in a biphasic pattern with an initial rapid rate of die-off followed by a period of slower die-off [19, 44]. Since we are simulating a single outbreak, waning immunity is ignored. See Appendix Section S4 for the full model equations, Table S1 for initial condition ranges, and Table 1 for parameter ranges.

## S3 Model Features

We incorporated different model features to examine mechanisms that can recreate the explosive outbreaks and low ARs characteristic of norovirus. We considered the following models (see Figure 1 for reference):

- **Baseline Model**: In this scenario, we simulated a fully susceptible population, with no individual exclusion. All individuals started in the susceptible, *S*, compartment.
- **Immunity Model**: In this scenario, we simulated partial population immunity with no individual exclusion. Because there is strain-dependent variation in the amount of protection innate resistance provides [32], and due to the fact that there is not an established correlate of norovirus protection that would be able to quantify acquired partial immunity, we examined different proportions of immunity. We assumed that those with innate resistance could not become infected at all and started as fully immune (in the *R* compartment), while those with acquired immunity started as partially immune (in *P*). Individuals in *P* could become infected, but not diseased. Non-diseased individuals were assumed to not be detectable during norovirus outbreaks and therefore were not counted in the numerator of the attack rate. Twenty percent of the population started in the *R* compartment i.e., with innate resistance [32], and we varied the total number with acquired immunity (*P*). We chose to vary the percentage starting with acquired immunity, because again there is not a well established correlate of protection [50]. Finally, we calibrated the proportion of individuals with acquired immunity to the data by sweeping over a broad range of Latin Hypercube Sampled [48] values (Table 1).
- **Individual Exclusion Model**: In this scenario, we simulated a fully susceptible population (i.e., all individuals started in the *S* compartment) with individual exclusion. During the simulation, a proportion of diseased individuals were removed from normal mixing and shedding, i.e., excluded. Excluded individuals do not contribute to transmission.
- **Combined Model**: In this scenario, we simulated partial population immunity, with individual exclusion.

Each of the above approaches were simulated stochastically. The stochastic simulation is a tau leaping version of the model [51] based on the Gillespie algorithm in which the stochastic model is approximate, but more efficient. The proportion of individuals across disease states is updated at each large predefined time step (the time interval is denoted *τ*). We then ran the model 10 times using different random number generator seeds for each parameter set and population size to account for stochastic variation.

## S4 Model Equations

As noted above, the model was simulated stochastically, but the equivalent ordinary differential equations are:

### Force of Infection

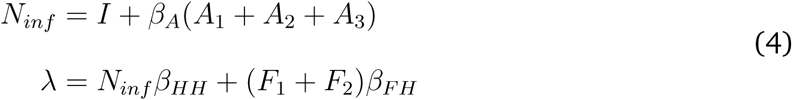

### Human Transmission Model

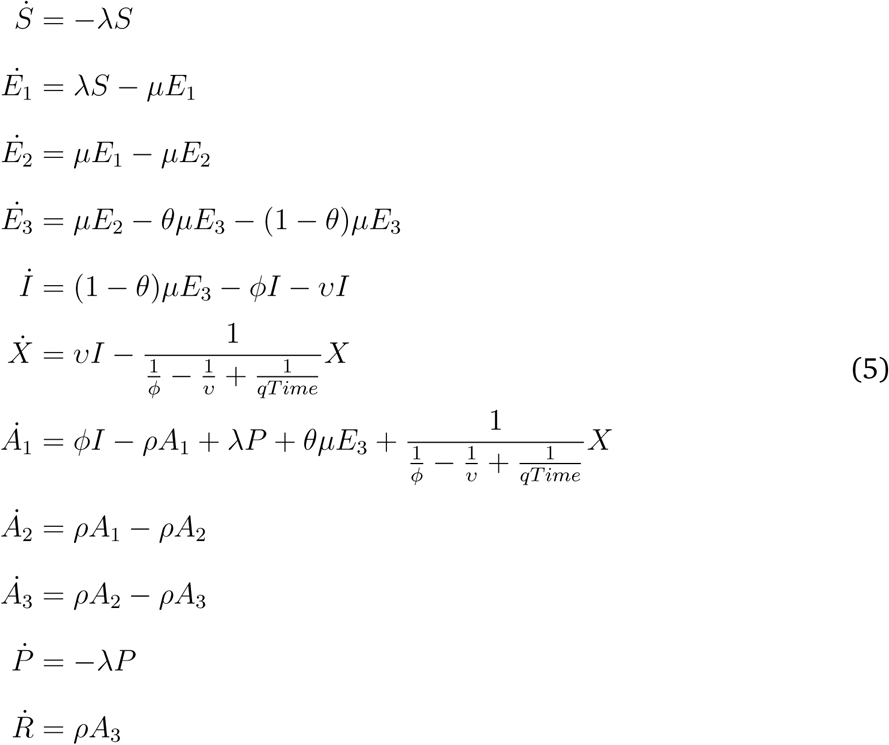

### Venue and Pathogens

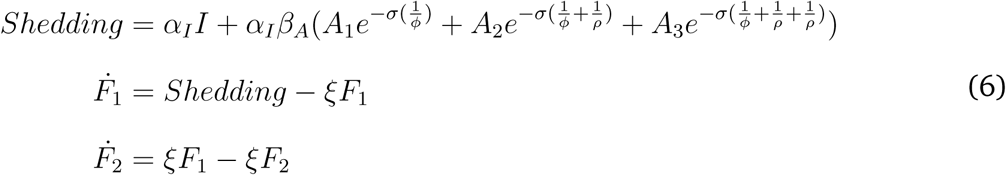

## S5 Initial Conditions

In the baseline model, we start all individuals as Susceptible (*S*). In the immunity and combined models, 20% of individuals start as fully recovered (*R*) and some proportion of individuals start with partial immunity (*P*). This proportion is randomly sampled between 0 and 80%. Finally, 10 million pathogens start in the *F*_1_ compartment to initiate the outbreak. However, this number was varied from 0 to 100 million in a sensitivity analysis. Another sensitivity analysis seeded the outbreak with a single infectious individual.

**Table S1:**
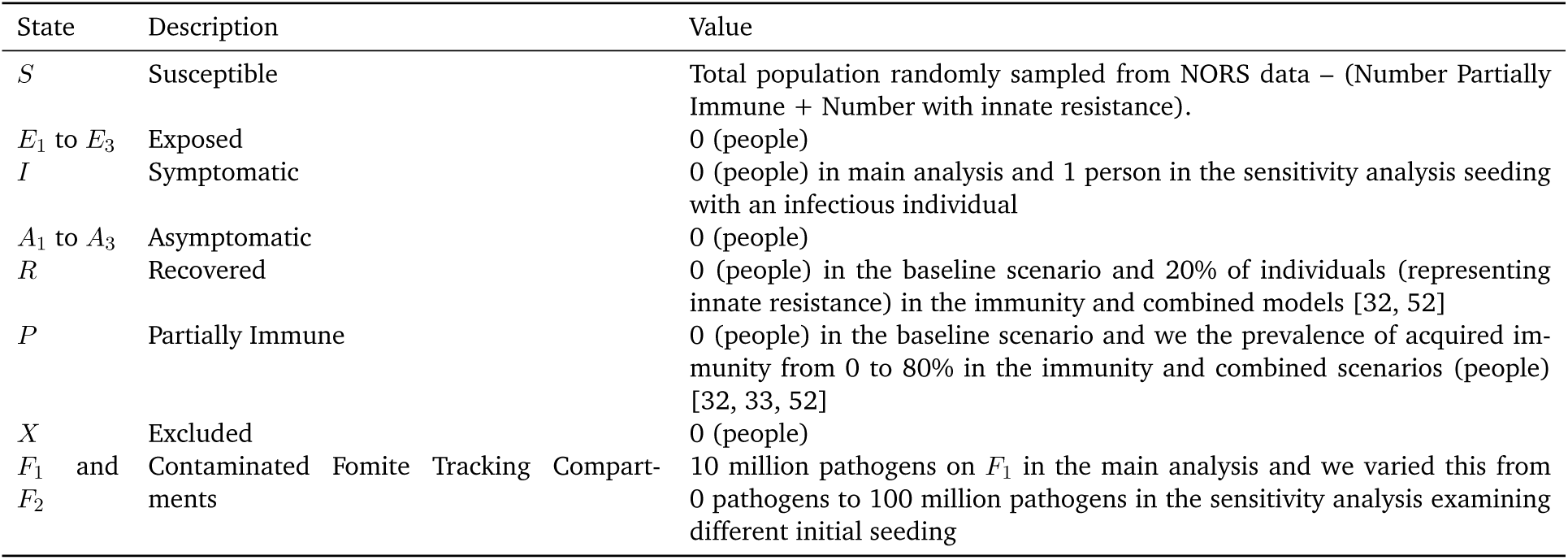
Initial Condition Values and Uncertainty Ranges

## S6 Calibration

We calibrated each venue–specific model separately to its corresponding NORS data using sample-importance-resampling [47]. This approach allowed us to obtain an array parameter sets upon which, if the model is run will recreate the NORS data distribution to the best ability of the model.

### S6.1 Initial Sample

We ran the model with 10,000 randomly sampled parameter and initial condition sets (collectively denoted ‘parameter sets’, and listed in Table 1) using Latin Hypercube Sampling [48]. Each parameter set was run in a school setting and separately, in a daycare setting. The only distinction between the model setup for schools and daycares was the starting population size (the sets of parameter values are the same), which is taken from the NORS outbreak data in the corresponding setting (Table S6). The NORS data includes separate attack rates (ARs) and populations for students and staff, but only one overall outbreak duration.

For a given venue–specific model, parameter sets were excluded from calibration if the outbreak was ongoing when the simulation ended (i.e., *>* 60 days, set according to the NORS data). We note that all the outbreaks in the NORS calibration dataset had ended by this point (the maximum duration was 40 days for daycare and 32 days for schools).

### S6.2 Calculating the Likelihood

We derived 2 kernel density estimates of the 3-dimensional probability distribution of the student ARs, student population size, and outbreak durations: one for NORS, *KDE*_*NORS*_, and one for the model results (generated based on our parameter ranges), *KDE*_*Model*_. The kernel density estimates were computed using the KS package in R which was designed for kernel smoothing of multidimensional data [53, 54]. We first derived the likelihood from the *KDE*_*NORS*_ value only and calibrated (fully informed prior). Next, because the pre-defined parameter ranges in the model represented our subjective best-guesses (i.e., our prior), we used *KDE*_*Model*_ (the model-derived kernel density estimate) to examine how weakening our prior (i.e., making it more uninformed) might affect results. Specifically, the likelihood estimate of a given parameter set was calculated by taking the NORS kernel density estimate value which corresponded to a given AR, population size, and outbreak duration from the model results and dividing that by the model kernel density estimate value i.e., we plugged our model outputs into both of these kernel density estimates, resulting in a fully uninformed (bounded uniform) prior. In other words, we looked up how likely our model results were to appear in the NORS data and divided by how likely our model results were in the entire pool of model results (wherein the shape of this pool of results is directly affected by our prior). By doing this, we effectively canceled the over-weighting of certain model results that occurred due to our parameter ranges. We subsequently investigated how results change when reducing the effect of the model kernel density estimate by using the following hill function:

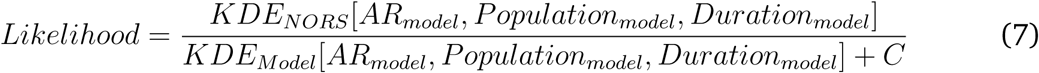

where [*AR*_*model*_, *Pop*_*model*_, *Duration*_*model*_] represent indices used to lookup the corresponding *KDE*_*NORS*_ value (i.e., to determine how likely our model results are to appear in the NORS data) and *KDE*_*model*_ value. *C* was the set to equal the 25^*th*^, 50^*th*^, 75^*th*^ percentiles of all *KDE*_*model*_ values to explore how weaker or stronger priors might affect calibration (i.e., as *C* increases the prior the effect of our predefined parameter ranges become stronger).

For each simulation, the AR was defined as the total number of symptomatic individuals divided by the total population and the outbreak duration was defined as the number of days from the first symptomatic incident case to the last symptomatic incident case.

### S6.3 Deriving our posterior

Finally, we resampled the parameter sets 2,500 times with replacement using the likelihoods as weights to obtain a final array of parameter sets that, if used as inputs for the model, could most closely recreate the NORS data distribution.

### S6.4 Calibration and Selecting a Prior

We initially calibrated using with a fully informed prior i.e., we looked up model outputs in *KDE*_*NORS*_ only (and did not divide by *KDE*_*Model*_). We found that although the models were generally able to recreate the distribution of the NORS data, the large majority of parameter sets yielded model results in specific regions of the NORS data distribution. Therefore, the majority of resampled parameter sets were taken from these regions and resulted in results (specifically outbreak durations) that were not consistent with the NORS data. See Appendix Section S7.1 for attack rates, durations, and Kullback-Leibler divergence from the fully informed prior analysis. The fact that model results were over-represented in specific regions was likely due to our pre-selected parameter ranges, therefore we explored whether making the prior weaker (i.e., plugging model outputs into *KDE*_*Model*_ and calculating likelihoods with the hill function see Appendix Table S7.1 for results) might improve calibration.

## S7 Selecting a Prior Results

We initially calibrated each model scenario to the NORS data without accounting for the distribution of model results. In other words, likelihoods were calculated based on how well the model results match the NORS data only. We next examined how weakening the prior might improve model calibration to NORS data.

Below are Kullback-Leibler divergence values for different values of *C* in the likelihood hill function (see Equation 7).

**Table S2:**
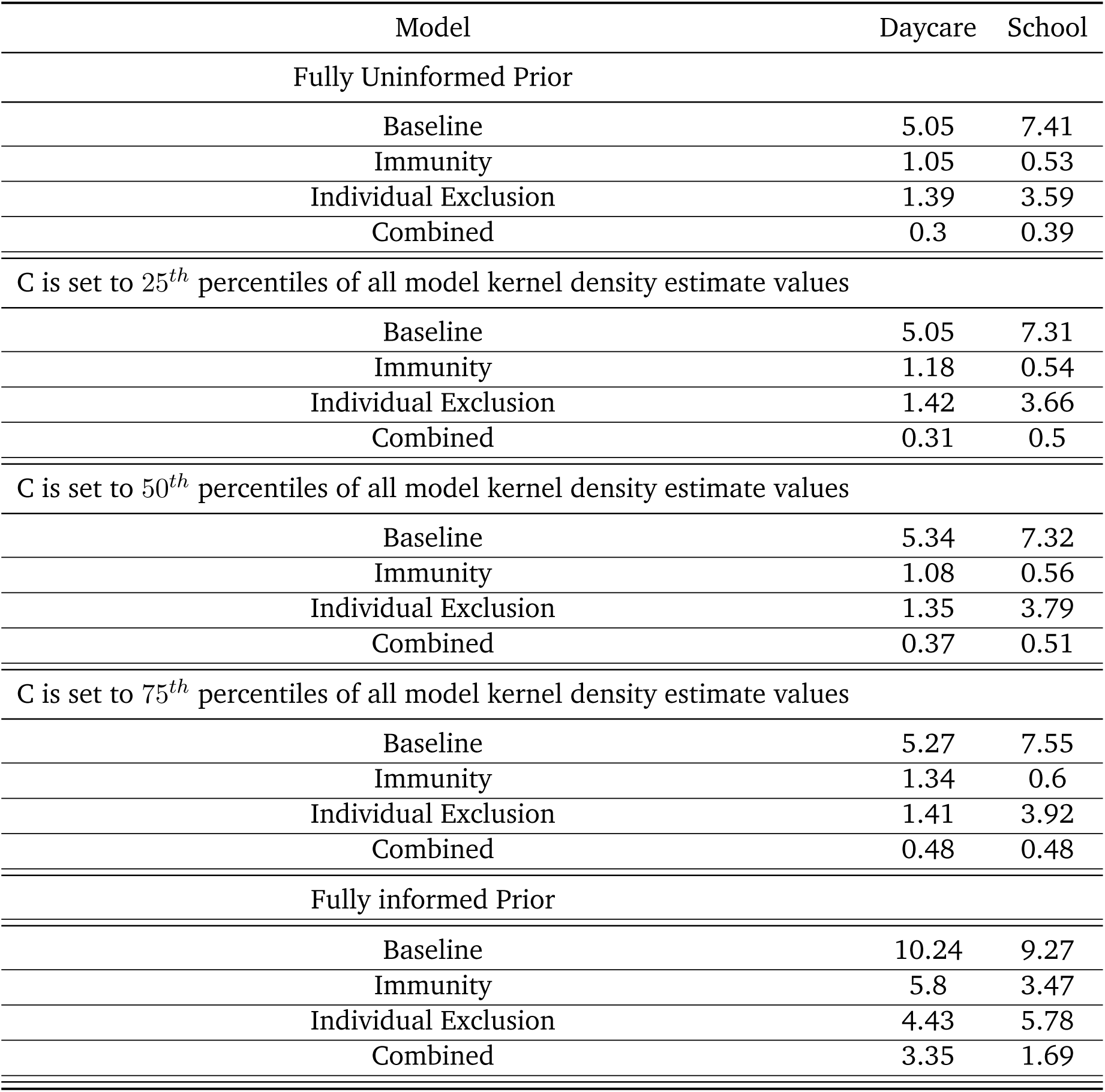
Venue-specific Kullback-Leibler divergence for All Models for different priors: Median (95% CI) [Mean]

### S7.1 Results from Fully Informed Prior Analysis

Below are the attack rates, durations, and Kullback-Leibler divergence from the fully informed prior analysis.

**Table S3:** Venue-specific Attack Rates for All Models (fully informed prior): Median (95% CI) [Mean]

| Model | Daycare | School |
| --- | --- | --- |
| Baseline | 66.2% (4%, 75.4%) [62.1%] | 64% (0%, 72.3%) [59.5%] |
| Immunity | 23.5% (3.1%, 50%) [23.7%] | 16.1% (2.2%, 53.5%) [20.1%] |
| Individual Exclusion | 61.9% (2.6%, 75%) [51.8%] | 58.3% (0%, 71.8%) [45.8%] |
| Combined | 20.9% (2.5%, 48.5%) [22.3%] | 15.3% (0.7%, 54.1%) [19.4%] |
| **NORS** | 21.6% (4.6%, 69.2%) [25.5%] | 15.3% (4.6%, 68.4%) [20.4%] |

**Table S4:**
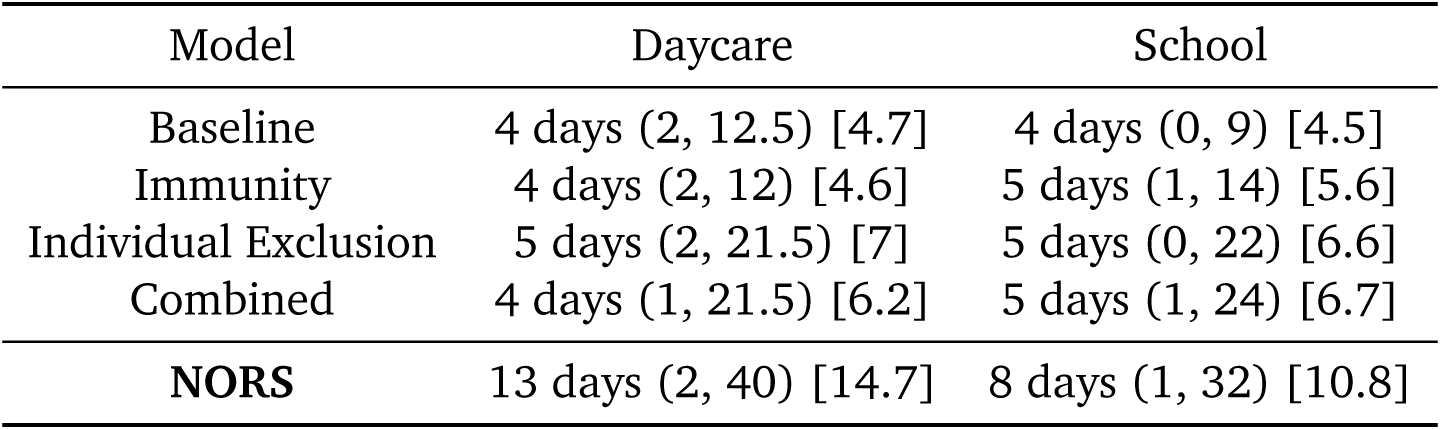
Venue-specific Outbreak Durations for All Models (fully informed prior): Median (95% CI) [Mean]

| Model | Daycare | School |
| --- | --- | --- |
| Baseline | 4 days (2, 12.5) [4.7] | 4 days (0, 9) [4.5] |
| Immunity | 4 days (2, 12) [4.6] | 5 days (1, 14) [5.6] |
| Individual Exclusion | 5 days (2, 21.5) [7] | 5 days (0, 22) [6.6] |
| Combined | 4 days (1, 21.5) [6.2] | 5 days (1, 24) [6.7] |
| **NORS** | 13 days (2, 40) [14.7] | 8 days (1, 32) [10.8] |

**Table S5:** Venue-specific Kullback-Leibler divergence for All Models (fully informed prior): Median (95% CI) [Mean]

| Model | Daycare | School |
| --- | --- | --- |
| Baseline | 10.24 | 9.27 |
| Immunity | 5.8 | 3.47 |
| Individual Exclusion | 4.43 | 5.78 |
| Combined | 3.35 | 1.69 |

Although the initial Latin Hypercube sampled simulations of the combined model were able to recreate almost the entire joint distribution of ARs and durations observed in the NORS data, a large fraction of the these simulations had very short outbreak durations (and these short duration simulations were weighted highly in the sample-importance-resampling as they are also common in NORS). This skewed the calibrated distributions of durations in the combined model toward the shorter-duration end of the NORS data, so that the median model-generated durations were lower than those in NORS. Therefore, for a model to be able to calibrate well, it is required to both (1) recreate the joint distribution of attack rates and durations in the NORS data, and (2) evenly distribute model runs across the distribution. This led us to examine how weaker priors accounting for the distribution of model results might improve the calibration. We found that the model scenarios consistently performed the approximately same relative to each other (e.g., the combined model always performed better than the baseline model)regardless of which prior we used (see Appendix Table S7.1). We ultimately chose to present results from a fully uninformed prior (see Equation 7 with *C*=0) because this yielded the best overall fit both graphically and with respect to Kullback-Leibler divergence.

## S8 NORS Calibration Ranges

We calibrated our models to the NORS data below. In total, there were 165 daycare outbreaks and 397 school outbreaks.

**Table S6:** Calibration Ranges from NORS Data

| Metric | Median (5<sup>th</sup> to 95<sup>th</sup> percentiles) [Mean] |
| --- | --- |
| Population sizes of daycare venue | 80 people (7, 410) [94.7] |
| Population sizes of school venue | 420 people (6, 6486) [447.3] |
| Attack rate within daycare venue | 21.5% (4.6%, 69.2%) [25.3%] |
| Attack rate within school venues | 15.3% (4.6%, 68.4%) [20.4%] |
| Outbreak duration within daycare venue | 13 days (2, 40) [14.6] |
| Outbreak duration within school venues | 8 days (1, 32) [10.8] |

## S9 Attack Rates vs. Outbreak Duration Stratified by NORS Population Sizes

**Figure S1:**
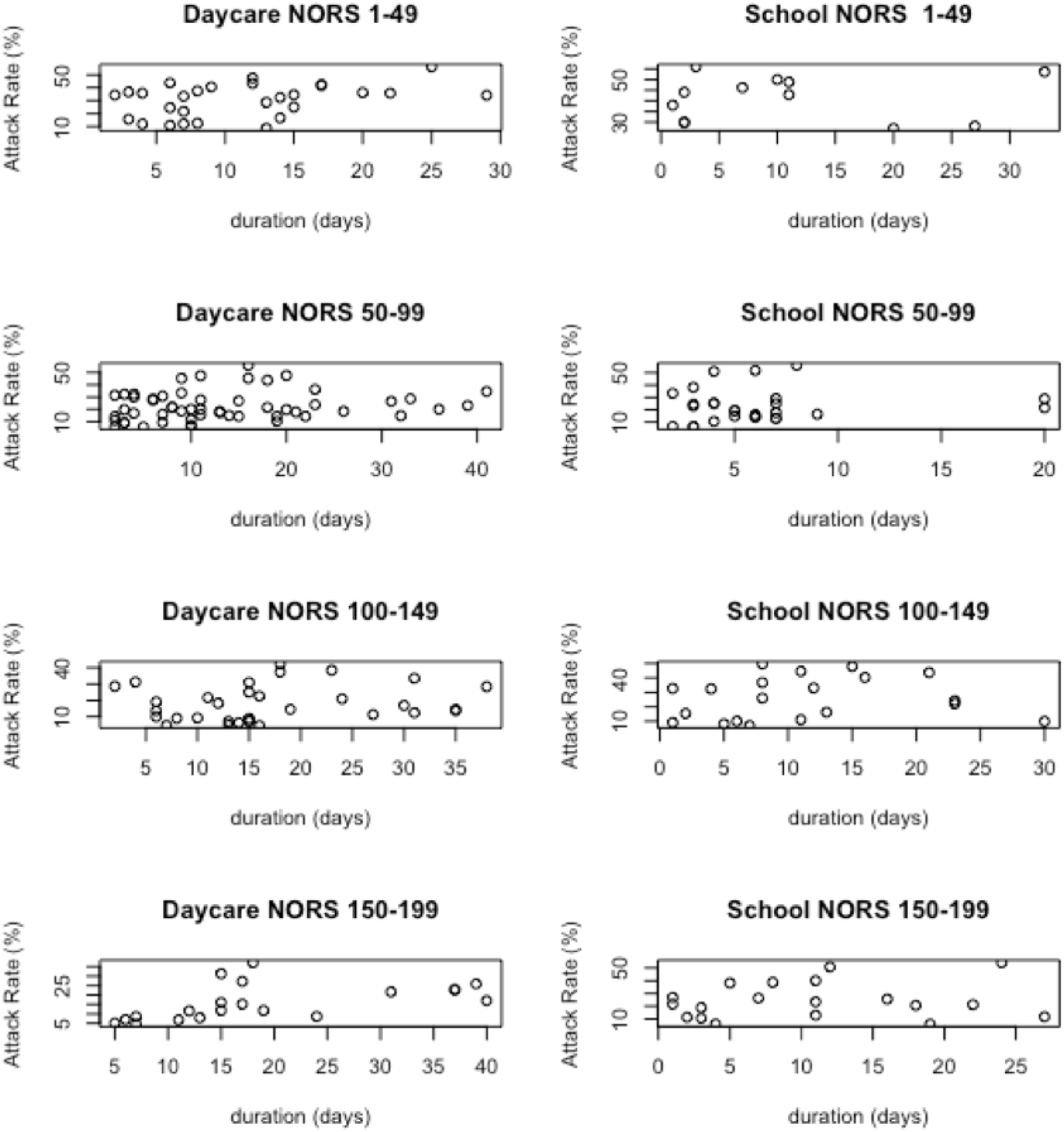
NORS data: Attack rates vs. outbreak duration stratified by exposed population size.

## S10 Results from Calibration for Each Model with NORS Data

Below are pairwise scatter plots examining joint distributions of attack rate (%), outbreak durations (days), and population sizes (people). The NORS data is in the upper left corner and all models are displayed with individual points colored by the log of the number of Times Calibrated. Points in white were not resampled.

## S11 Sensitivity Analyses Related to Initial Conditions

We conducted sensitivity analyses to ensure that our model results were robust to key simplifying assumptions. To assess whether the outbreak durations were affected by the choice of initial conditions in different compartments, we conducted two sensitivity analyses. First, we ran the model varying the number of pathogens starting in the environment from 0 to 100 million and second, we seeded the model with a single infectious individual.

**Figure S2:**
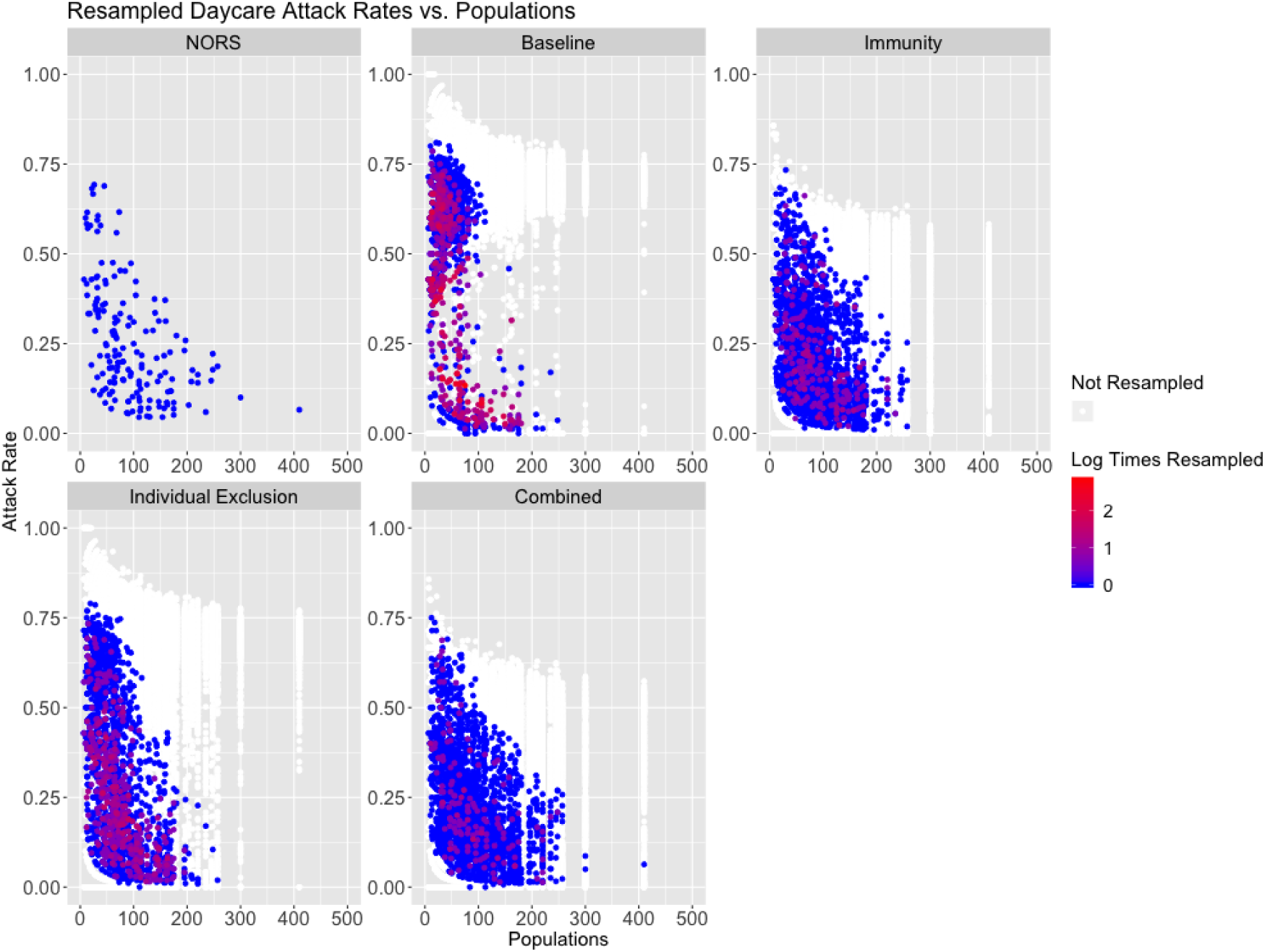
Attack rate vs. population in daycares for the NORS data (top left), with all remaining panels showing results from resampled parameter and initial conditions by model scenario. Points correspond to individual parameter sets and are colored by the amount of times they were resampled.

**Figure S3:**
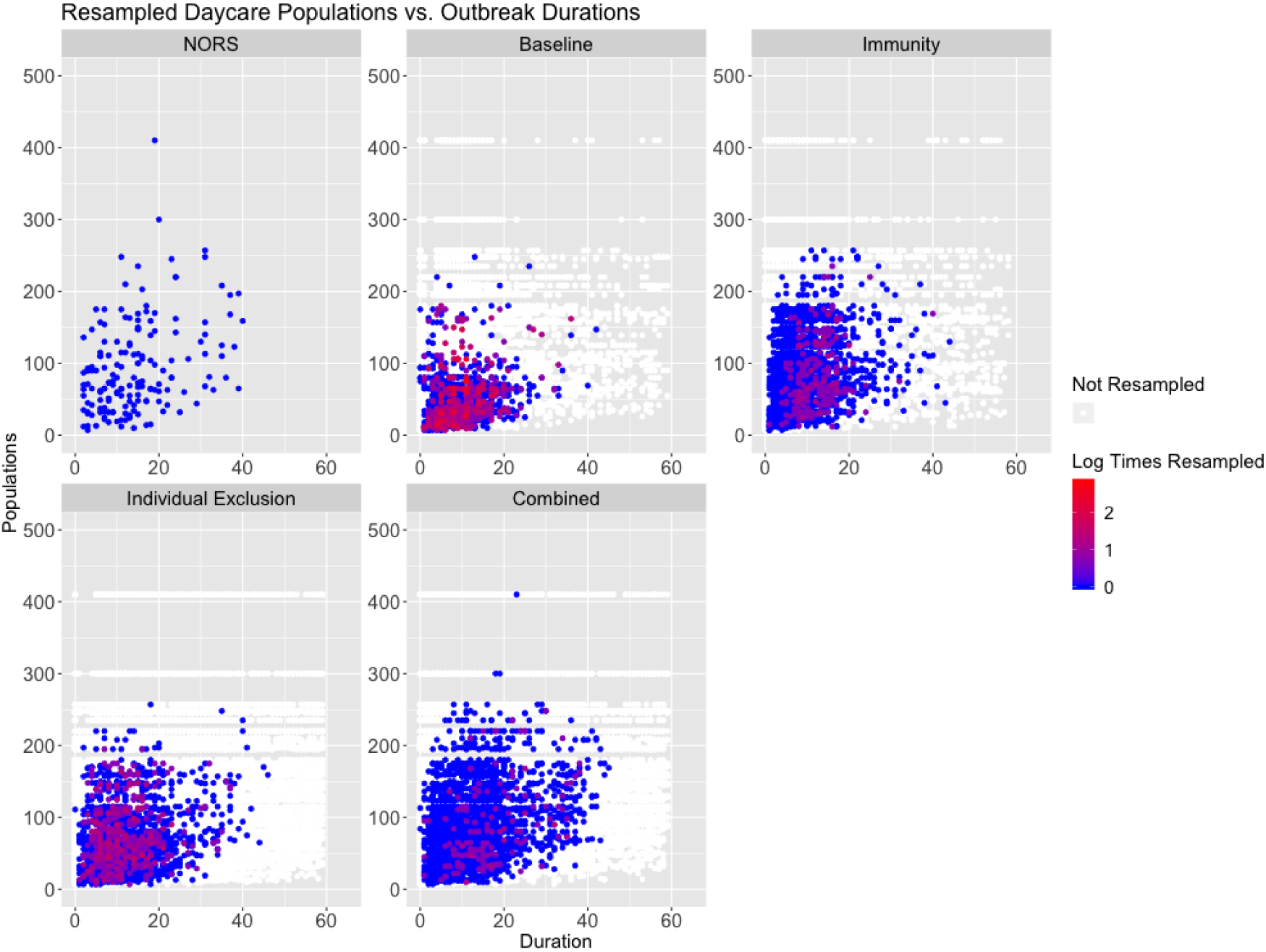
Population vs. outbreak duration in daycares for the NORS data (top left), with all remaining panels showing results from resampled parameter and initial conditions by model scenario. Points correspond to individual parameter sets and are colored by the amount of times they were resampled.

**Figure S4:**
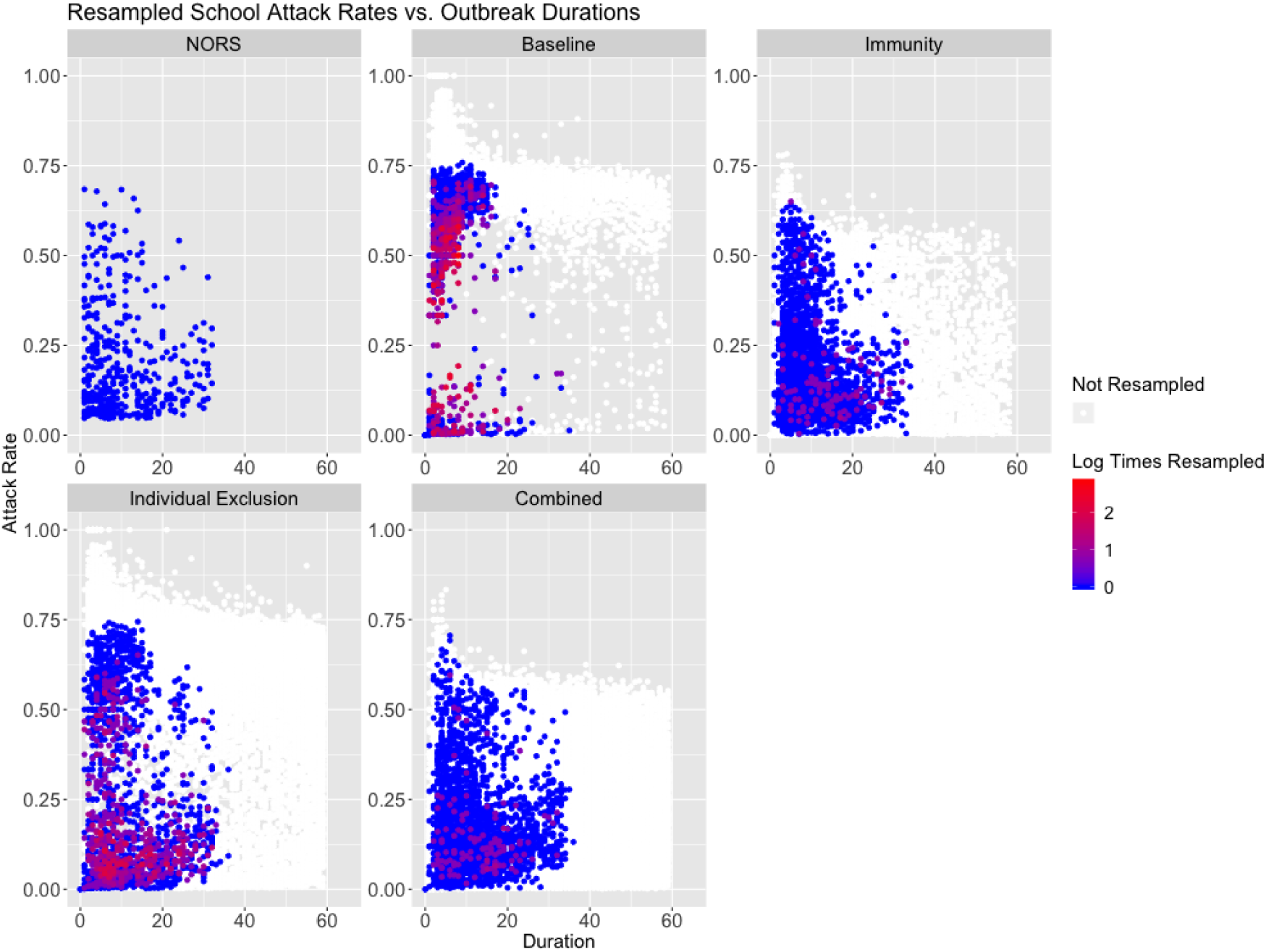
Attack rate vs. outbreak duration in schools for the NORS data (top left), with all remaining panels showing results from resampled parameter and initial conditions by model scenario. Points correspond to individual parameter sets and are colored by the amount of times they were resampled.

**Figure S5:**
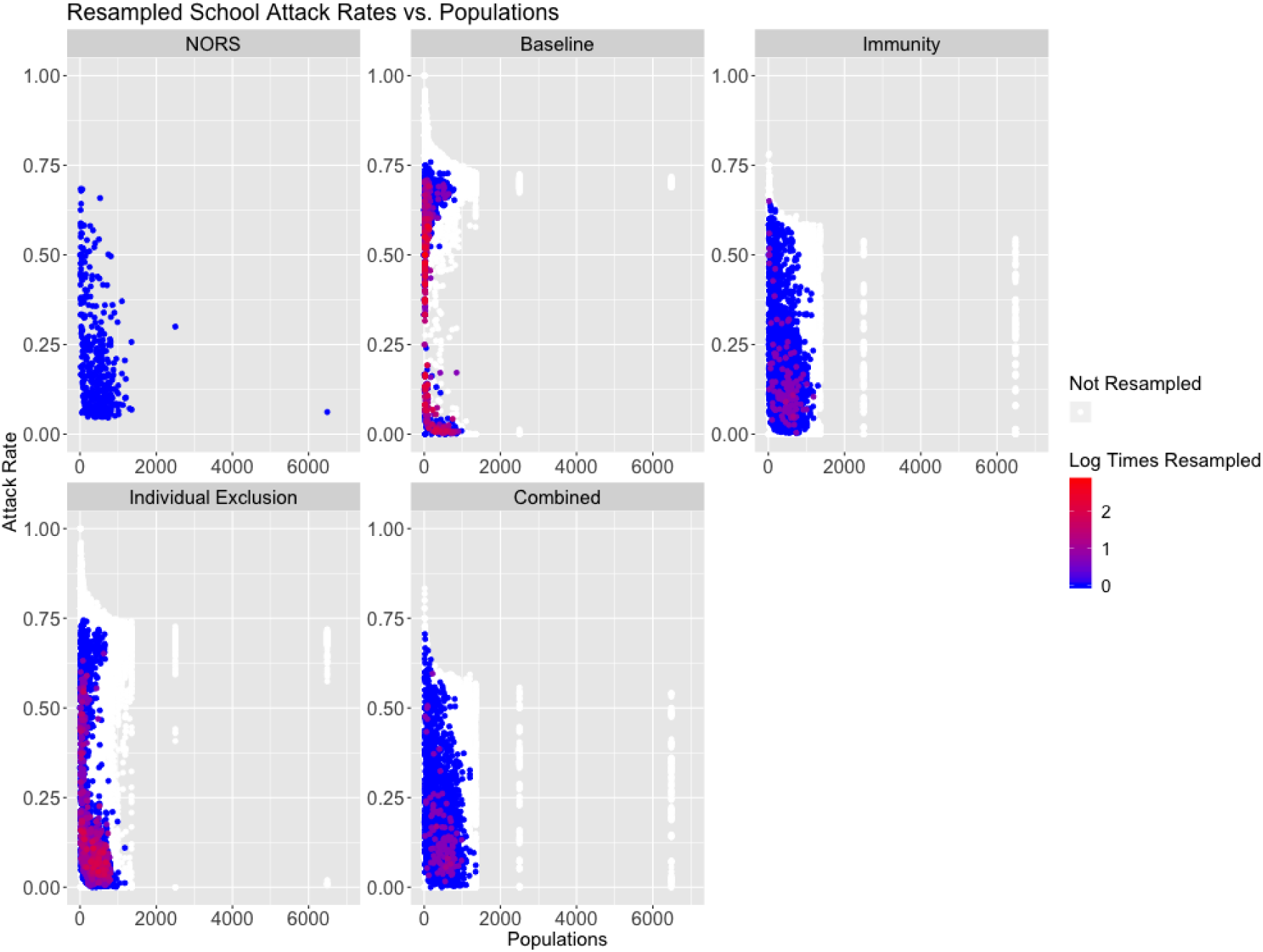
Attack rate vs. population in schools for the NORS data (top left), with all remaining panels showing results from resampled parameter and initial conditions by model scenario. Points correspond to individual parameter sets and are colored by the amount of times they were resampled.

**Figure S6:**
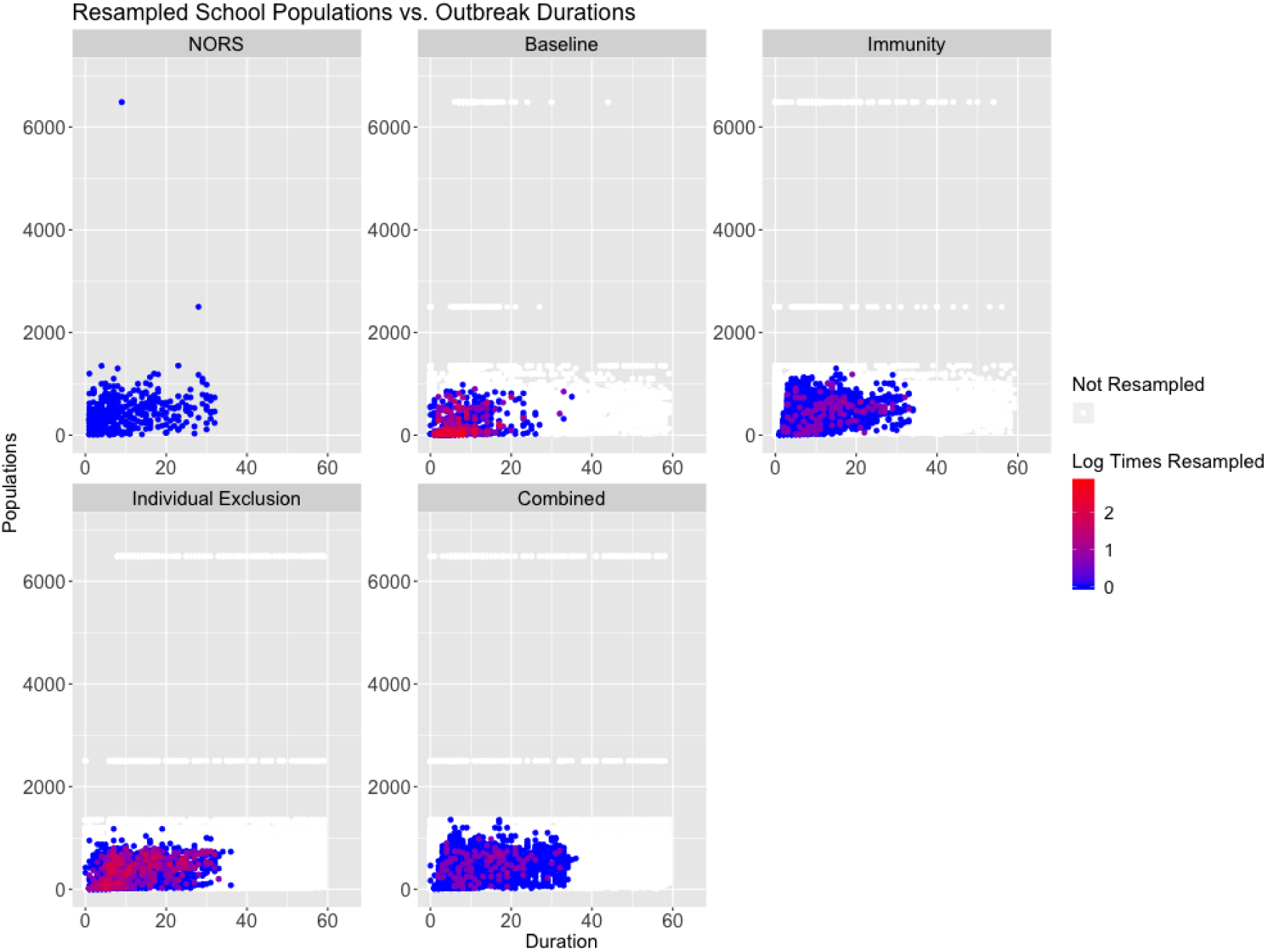
Population vs. outbreak duration in schools for the NORS data (top left), with all remaining panels showing results from resampled parameter and initial conditions by model scenario. Points correspond to individual parameter sets and are colored by the amount of times they were resampled.

### S11.1 Sensitivity Analysis Results

According to the Kullback-Leibler divergence, all sensitivity analyses performed fairly well and were relatively close to the original combined model. Overall, the combined was lower according to Kullback-Leibler divergence in the daycare model, but seeding varying pathogens in the the environment had a lower Kullback-Leibler divergence in the school model. See Table 2 for Kullback-Leibler divergence values of the main analyses.

Below are attack rates and durations from seeding scenario the sensitivity analyses.

**Table S7:** Venue-specific Attack Rates for Sensitivity Analyses: Median (95% CI) [Mean]

| Model | Daycare | School |
| --- | --- | --- |
| Seeding: Varying Pathogens in Environment | 19.9% (3.4%, 51.5%) [22.4%] | 14.8% (2.1%, 53.7%) [19.1%] |
| Seeding: Diseased Individual | 21.1% (3.3%, 56.4%) [23.9%] | 15.4% (2.1%, 55%) [19.4%] |
| **NORS** | 21.5% (4.6%, 69.2%) [25.3%] | 15.3% (4.6%, 68.4%) [20.4%] |

**Table S8:**
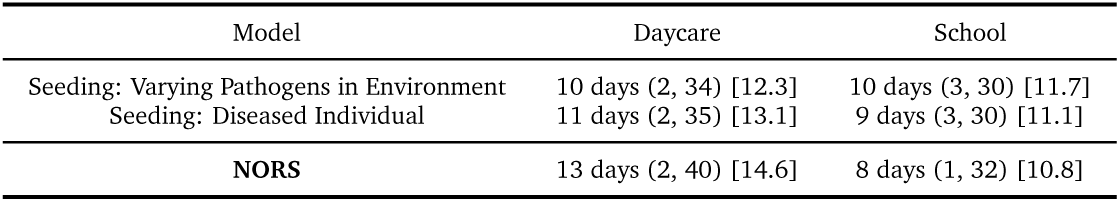
Venue-specific Outbreak Durations for Sensitivity Analyses: Median (95% CI) [Mean]

### S12 Staff and Students Model

We added staff into the model, to understand whether or not they can affect how norovirus is spread within venues.

### S12.1 Students and Staff Model for Daycare and School

In the staff and student model, there is a staff age group and a student age group. To derive the human-to-human transmission rates, we assume that the younger age group (i.e., the students) transmit at higher rates than the older age group (i.e., the staff) due to both contact rates [55] and susceptibility decreasing with age (e.g. represented by levels of norovirus antibody titers [56]). Specifically, the human-to-human transmission matrix is derived by taking the *β*_*HH*_ from the students only model and setting that to the student to student transmission rate. Next, we assume that the inter-age transmission rates (i.e., staff to student and student to staff transmission are equal) and calculate that by multiplying the student to student transmission rate by a randomly sampled reduction factor between [0,1]. Finally, the staff-to-staff transmission rate is calculated by multiplying the inter-age transmission rate by a randomly sampled reduction factor between [0,1] (this factor is also used to derive the fomite-to-staff transmission rate).

Next, for fomite-to-human transmission there are two rates, one for students and one for staff. The fomite-to-student transmission rate is calculated in the same way as the student only model i.e., *β*_*HH*_ multiplied by a randomly sampled parameter between [0, 2]. The fomite-to-staff transmission rate is derived by multiplying the fomite-to-student rate by the same factor used to derive the staff-to-staff transmission rate (mentioned above) between [0,1]. Overall, the force of infection for the staff and students model is as follows:

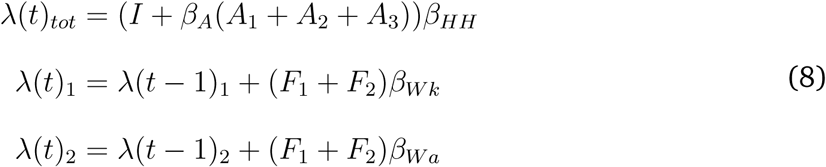

where *λ*(*t*)_*tot*_ is the total force of infection (and is a vector representing the student force of infection as the first element and the staff force of infection as the second element. Thus the human-to-human transmission rate (*β*_*HH*_) is a 2 by 2 matrix. *λ*(*t*)_1_ and *λ*(*t*)_2_ are added to the force of infection for students and staff, respectively. Finally, *β*_*W k*_ and *β*_*W a*_ are the fomite-to-human transmission rates for students and staff, respectively.

Finally, with respect to shedding, staff and students shed into a single shared environment. Thus, the shedding and fomite tracking equations are as follows:

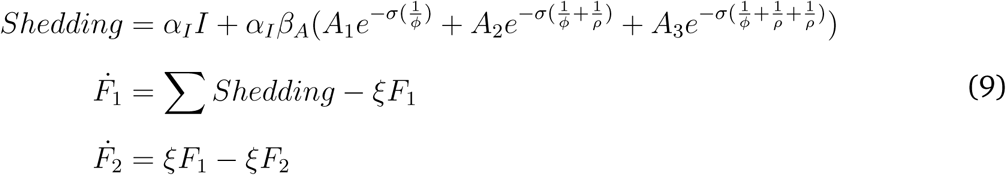

where the sum of shedding across both age groups is added to the *F*_1_ compartment because there is a single environmental compartment in each venue.

For the immunity and combined models we assumed that staff had higher rates of partial immunity than children [56].

All other model equations are the same as the student only model, we just vectorized the equations to keep track of student and staff compartments separately. see Appendix Section S4 for details.

### S12.2 Students and Staff Model Likelihood Calculation

To derive an overall likelihood for a given venue, we took the NORS kernel density estimate values which corresponded to a given AR and population size for students from the model and divided by the model kernel density estimate values which corresponded to a given AR and population size for students from the model. We multiplied this by the corresponding staff value (i.e., NORS kernel density estimate divided by model kernel density estimate), and finally, multiplied by the NORS kernel density estimate value divided by the model kernel density estimate value which corresponded to a given outbreak duration and total venue population from the model. More details can be found in Section 5.3. We did not calculate a full 5-dimensional kernel density estimate due to computational limitations. Therefore, a key limitation in our approach for this sensitivity analysis is that we are assuming independence between different kernel density estimates (e.g., the student joint distribution of ARs and population size is assumed to be independent of the staff distribution).

We calibrated the student and staff model to 137 daycare and 240 school outbreaks.

### S12.3 Students and Staff Model Results

According to the Kullback-Leibler divergence, the combined model calibrated best to the NORS data. Although the median results were closer to NORS data for model scenarios, the attack rate ranges were wider for the immunity model, and the outbreak duration ranges were wider for the individual exclusion model.

**Table S9:** Kullback-Leibler Divergence Metrics for the Students and Staff Model

| Model | Daycare Students | Daycare Staff | Daycare Outbreak Duration | School Students | School Staff | School Outbreak Duration |
| --- | --- | --- | --- | --- | --- | --- |
| Baseline | 13.75 | 15.41 | 7.94 | 12.73 | 12.74 | 6.77 |
| Immunity | 0.28 | 0.79 | 0.25 | 0.33 | 0.89 | 0.27 |
| Individual Exclusion 0.62 | 2.56 | 0.86 | 4.43 | 2.81 | 2.02 |  |
| **Combined** | **0.32** | **0.53** | **0.14** | **0.12** | **0.33** | **0.15** |

**Table S10:**
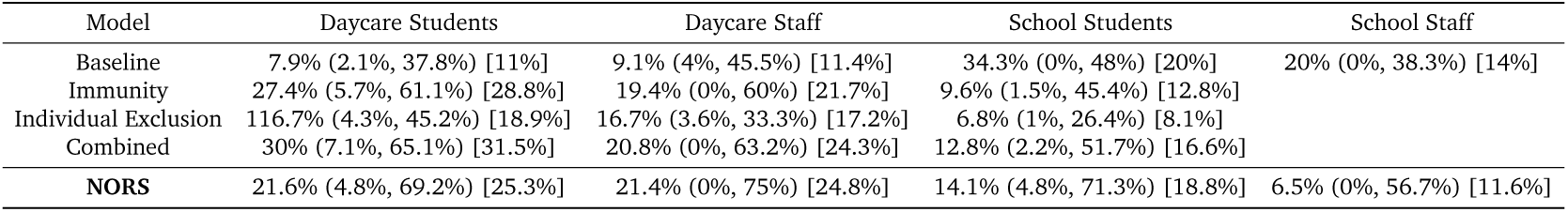
Venue-specific Attack Rates for Students and Staff Model: Median (95% CI) [Mean]

**Table S11:** Venue-specific Outbreak Durations for Students and Staff Model: Median (95% CI) [Mean]

| Model | Daycare | School |
| --- | --- | --- |
| Baseline | 14 days (2, 30) [15.1] | 12 days (1, 30) [10.9] |
| Immunity | 14 days (4, 35) [15.2] | 14 days (3, 33) [14.9] |
| Individual Exclusion | 24 days (4, 43) [24.6] | 20 days (3, 33) [20.2] |
| Combined | 15 days (4, 39) [16.9] | 13 days (3, 33) [14.4] |
| **NORS** | 14 days (2, 40) [15.5] | 11 days (2, 33) [12.9] |

## Notes

### Competing Interest Statement

Dr. Lopman reports personal fees from Takeda Pharmaceuticals, CDC Foundation, and Hall Booth Smith, P.C. outside the submitted work.

### Funding Statement

This Study was funded by the Joint Initiative for Vaccine Economics, Phase 5, a cooperative agreement between the University of Michigan and the Centers for Disease Control and Prevention (U01IP000965). Additionally, this study was funded by the National Institute of General Medical Sciences (U01GM110712).

## References

[1] Hall AJ, Lopman BA, Payne DC, Patel MM, Gastañaduy PA, Vinjé J, et al. Norovirus disease in the United States. Emerging infectious diseases. 2013;19(8):1198.

[2] Steele MK, Remais JV, Gambhir M, Glasser JW, Handel A, Parashar UD, et al. Targeting pediatric versus elderly populations for norovirus vaccines: a model-based analysis of mass vaccination options. Epidemics. 2016;17:42–49.

[3] Heun EM, Vogt RL, Hudson PJ, Parren S, Gary GW. Risk factors for secondary transmission in households after a common-source outbreak of Norwalk gastroenteritis. American journal of epidemiology. 1987;126(6):1181–1186.

[4] Marks P, Vipond I, Regan F, Wedgwood K, Fey R, Caul E. A school outbreak of Norwalk-like virus: evidence for airborne transmission. Epidemiology & Infection. 2003;131(1):727–736.

[5] Götz H, Ekdahl K, Lindbäck J, de Jong B, Hedlund KO, Giesecke J. Clinical spectrum and transmission characteristics of infection with Norwalk-like virus: findings from a large community outbreak in Sweden. Clinical Infectious Diseases. 2001;33(5):622–628.

[6] Hoebe CJ, Vennema H, de Roda Husman AM, van Duynhoven YT. Norovirus outbreak among primary schoolchildren who had played in a recreational water fountain. The Journal of infectious diseases. 2004;p. 699–705.

[7] Isakbaeva ET, Bulens SN, Beard RS, Adams S, Monroe SS, Chaves SS, et al. Norovirus and child care: challenges in outbreak control. The Pediatric infectious disease journal. 2005;24(6):561–563.

[8] Morioka S, Sakata T, Tamaki A, Shioji T, Funaki A, Yamamoto Y, et al. A food-borne norovirus outbreak at a primary school in Wakayama Prefecture. Japanese journal of infectious diseases. 2006;59(3):205.

[9] Marsh Z, Grytdal S, Beggs J, Leshem E, Gastañaduy P, Rha B, et al. The unwel-come houseguest: secondary household transmission of norovirus. Epidemiology & Infection. 2018;146(2):159–167.

[10] Adler JL, Zickl R. Winter vomiting disease. The Journal of infectious diseases. 1969;119(6):668–673.

[11] Wikswo ME, Hall AJ. Outbreaks of acute gastroenteritis transmitted by person-to-person contact—United States, 2009–2010. Morbidity and Mortality Weekly Report: Surveillance Summaries. 2012;61(9):1–12.

[12] Verhoef L, Hewitt J, Barclay L, Ahmed S, Lake R, Hall AJ, et al. Norovirus Genotype Profiles Associated with Foodborne Transmission, 1999 ?” 2012. Emerging infectious diseases. 2015;21(4):592.

[13] Nygård K, Torvén M, Ancker C, Knauth SB, Hedlund KO, Giesecke J, et al. Emerging genotype (GGIIb) of norovirus in drinking water, Sweden. Emerging infectious diseases. 2003;9(12):1548.

[14] Jones EL, Kramer A, Gaither M, Gerba CP. Role of fomite contamination during an outbreak of norovirus on houseboats. International journal of environmental health research. 2007;17(2):123–131.

[15] Cheesbrough J, Green J, Gallimore C, Wright P, Brown D. Widespread environmental contamination with Norwalk-like viruses (NLV) detected in a prolonged hotel outbreak of gastroenteritis. Epidemiology & Infection. 2000;125(1):93–98.

[16] Evans MR, Meldrum R, Lane W, Gardner D, Ribeiro C, Gallimore C, et al. An outbreak of viral gastroenteritis following environmental contamination at a concert hall. Epidemiology & Infection. 2002;129(2):355–360.

[17] Lopman B, Gastanaduy P, Park GW, Hall AJ, Parashar UD, Vinjé J. Environmental transmission of norovirus gastroenteritis. Current opinion in virology. 2012;2(1):96–102.

[18] Milbrath M, Spicknall I, Zelner J, Moe C, Eisenberg J. Heterogeneity in norovirus shedding duration affects community risk. Epidemiology & Infection. 2013;141(8):1572–1584.

[19] Fallahi S, Mattison K. Evaluation of murine norovirus persistence in environments relevant to food production and processing. Journal of food protection. 2011;74(11):1847–1851.

[20] D’Souza DH, Sair A, Williams K, Papafragkou E, Jean J, Moore C, et al. Persistence of caliciviruses on environmental surfaces and their transfer to food. International journal of food microbiology. 2006;108(1):84–91.

[21] Cheesbrough J, Barkess-Jones L, Brown D. Possible prolonged environmental survival of small round structured viruses. Journal of Hospital Infection. 1997;35(4):325–326.

[22] Clay S, Maherchandani S, Malik YS, Goyal SM. Survival on uncommon fomites of feline calicivirus, a surrogate of noroviruses. American journal of infection control. 2006;34(1):41–43.

[23] Teunis P, Sukhrie F, Vennema H, Bogerman J, Beersma M, Koopmans M. Shedding of norovirus in symptomatic and asymptomatic infections. Epidemiology & Infection. 2015;143(8):1710–1717.

[24] Atmar RL, Opekun AR, Gilger MA, Estes MK, Crawford SE, Neill FH, et al. Determination of the 50% human infectious dose for Norwalk virus. The Journal of infectious diseases. 2013;209(7):1016–1022.

[25] Parrino TA, Schreiber DS, Trier JS, Kapikian AZ, Blacklow NR. Clinical immunity in acute gastroenteritis caused by Norwalk agent. New England Journal of Medicine. 1977;297(2):86–89.

[26] Johnson PC, Mathewson JJ, DuPont HL, Greenberg HB. Multiple-challenge study of host susceptibility to Norwalk gastroenteritis in US adults. Journal of Infectious Diseases. 1990;161(1):18–21.

[27] Karst SM. Pathogenesis of noroviruses, emerging RNA viruses. Viruses. 2010;2(3):748–781.

[28] Xu H, Lin Q, Chen C, Zhang J, Zhang H, Hao C. Epidemiology of norovirus gastroenteritis outbreaks in two primary schools in a city in eastern China. American journal of infection control. 2013;41(10):e107–e109.

[29] Paunio M, Peltola H, Valle M, Davidkin I, Virtanen M, Heinonen OP. Explosive school-based measles outbreak: intense exposure may have resulted in high risk, even among revaccinees. American journal of epidemiology. 1998;148(11):1103–1110.

[30] Wells WF, Wells MW, Wilder TS, et al. The Environmental Control of Epidemic Contagion. I. An Epidemiologic Study of Radiant Disinfection of Air in Day Schools. American Journal of Hygiene. 1942;35(1):97–121.

[31] Hall AJ, Wikswo ME, Manikonda K, Roberts VA, Yoder JS, Gould LH. Acute gastroenteritis surveillance through the national outbreak reporting system, United States. Emerging infectious diseases. 2013;19(8):1305.

[32] Currier RL, Payne DC, Staat MA, Selvarangan R, Shirley SH, Halasa N, et al. Innate susceptibility to norovirus infections influenced by FUT2 genotype in a United States pediatric population. Clinical Infectious Diseases. 2015;60(11):1631–1638.

[33] Nurminen K, Blazevic V, Huhti L, Räsänen S, Koho T, Hytönen VP, et al. Prevalence of norovirus GII-4 antibodies in Finnish children. Journal of medical virology. 2011;83(3):525–531.

[34] Kulkarni R, Lole K, Chitambar SD. Seroprevalence of antibodies against GII. 4 norovirus among children in Pune, India. Journal of medical virology. 2016;88(9):1636–1640.

[35] Melhem NM. Norovirus vaccines: correlates of protection, challenges and limitations. Human vaccines & immunotherapeutics. 2016;12(7):1653–1669.

[36] Hall AJ, Vinjé J, Lopman B, Park GW, Yen C, Gregoricus N, et al. Updated norovirus outbreak management and disease prevention guidelines. Morbidity and Mortality Weekly Report: Recommendations and Reports. 2011;60(3):1–15.

[37] Barclay L, Park G, Vega E, Hall A, Parashar U, Vinjé J, et al. Infection control for norovirus. Clinical microbiology and infection. 2014;20(8):731–740.

[38] Keeling MJ, Ross JV. On methods for studying stochastic disease dynamics. Journal of The Royal Society Interface. 2008;5(19):171–181.

[39] NORS;. Accessed: 2018-08-23. https://www.cdc.gov/NORS/about.html.

[40] NORS;. Accessed: 2018-11-23. https://www.cdc.gov/nors/downloads/guidance

[41] R Core Team. R: A Language and Environment for Statistical Computing. Vienna, Austria; 2016. Available from: https://www.R-project.org/pdf.

[42] Lee RM, Lessler J, Lee RA, Rudolph KE, Reich NG, Perl TM, et al. Incubation periods of viral gastroenteritis: a systematic review. BMC infectious diseases. 2013;13(1):446.

[43] Atmar RL, Opekun AR, Gilger MA, Estes MK, Crawford SE, Neill FH, et al. Norwalk virus shedding after experimental human infection. Emerging infectious diseases. 2008;14(10):1553.

[44] Verhaelen K, Bouwknegt M, Lodder-Verschoor F, Rutjes SA, de Roda Husman AM. Persistence of human norovirus GII. 4 and GI. 4, murine norovirus, and human adenovirus on soft berries as compared with PBS at commonly applied storage conditions. International journal of food microbiology. 2012;160(2):137–144.

[45] Rockx B, de Wit M, Vennema H, Vinjé J, de Bruin E, van Duynhoven Y, et al. Natural history of human calicivirus infection: a prospective cohort study. Clinical Infectious Diseases. 2002;35(3):246–253.

[46] Gaythorpe K, Trotter CL, Lopman B, Steele M, Conlan A. Norovirus transmission dynamics: a modelling review. Epidemiology & Infection. 2018;146(2):147–158.

[47] Rubin DB. Using the SIR algorithm to simulate posterior distributions. Bayesian statistics. 1988;3:395–402.

[48] Stein M. Large sample properties of simulations using Latin hypercube sampling. Technometrics. 1987;29(2):143–151.

[49] Kullback S, Leibler RA. On information and sufficiency. The annals of mathematical statistics. 1951;22(1):79–86.

[50] Ramani S, Estes MK, Atmar RL. Correlates of protection against norovirus infection and disease—where are we now, where do we go? PLoS pathogens. 2016;12(4):e1005334.

[51] Gillespie DT. Approximate accelerated stochastic simulation of chemically reacting systems. The Journal of Chemical Physics. 2001;115(4):1716–1733.

[52] Lindesmith L, Moe C, Marionneau S, Ruvoen N, Jiang X, Lindblad L, et al. Human susceptibility and resistance to Norwalk virus infection. Nature medicine. 2003;9(5):548.

[53] Deng H, Wickham H. Density estimation in R. Electronic publication. 2011;.

[54] Duong T, et al. ks: Kernel density estimation and kernel discriminant analysis for multivariate data in R. Journal of Statistical Software. 2007;21(7):1–16.

[55] Mossong J, Hens N, Jit M, Beutels P, Auranen K, Mikolajczyk R, et al. Social contacts and mixing patterns relevant to the spread of infectious diseases. PLoS medicine. 2008;5(3):e74.

[56] Carmona-Vicente N, Fernández-Jiménez M, Ribes JM, Téllez-Castillo CJ, KhodayarPardo P, Rodríguez-Diaz J, et al. Norovirus infections and seroprevalence of genotype gii. 4-specific antibodies in a spanish population. Journal of medical virology. 2015;87(4):675–682.

